# The role of etiology in the identification of clinical markers of consciousness: comparing EEG alpha power, complexity, and spectral exponent

**DOI:** 10.1101/2024.03.20.24304639

**Authors:** Charlotte Maschke, Laouen Belloli, Dragana Manasova, Jacobo D. Sitt, Stefanie Blain-Moraes

## Abstract

In the search for EEG markers of human consciousness, alpha power has long been considered a reliable marker which is fundamental for the assessment of unresponsive patients from all etiologies. However, recent evidence questioned the role of alpha power as a marker of consciousness and proposed the spectral exponent and spatial gradient as more robust and generalizable indexes. In this study, we analyzed a large-scale dataset of 260 unresponsive patients and investigated etiology-specific markers of level of consciousness, responsiveness and capacity to recover. We compare a set of candidate EEG makers: 1) absolute, relative and flattened alpha power; 2) spatial ratios; 3) the spectral exponent; and 4) signal complexity. Our results support the claim that alpha power is an etiology-specific marker, which has higher diagnostic value for anoxic patients. Meanwhile, the spectral slope showed diagnostic value for non-anoxic patients only. Changes in relative power and signal complexity were largely attributable to changes in the spectral slope. Grouping unresponsive patients from different etiologies together can confound or obscure the diagnostic value of different EEG markers of consciousness. Our study highlights the importance of analyzing different etiologies independently and emphasizes the need to develop clinical markers which better account for inter-individual and etiology-dependent differences.

## 2 Introduction

For unresponsive brain-injured patients, the evaluation of level of consciousness and capacity to recover underpins the most crucial decisions about their treatment and continuation of care. The quest for neurophysiological markers of (un)consciousness plays a fundamental role in the development of clinical tools and the understanding of the neurophysiological process underlying consciousness.

The loss of oscillatory power – mainly in the alpha frequency band – has often been described as the principal marker for detecting loss of consciousness (Chennu et al. 2014; Sitt et al. 2014; Naro et al. 2016; Piarulli et al. 2016). However, recent work by Colombo et al. (2023) raised strong concerns about the role of alpha power as a marker of consciousness. Instead of marking loss of consciousness, Colombo et al (2023) attributed loss of alpha power to the suppression of cortical activity, which is specific to severe postanoxic injury. As an alternative marker, Colombo et al (2023) proposed the use of 1) the spectral exponent and 2) the alpha posterior anterior ratio. In contrast to alpha power, both measures stratified levels of consciousness in non-anoxic patients and generalized well to pharmacological-induced unconsciousness (Colombo et al. 2023).

Indeed, the electroencephalogram (EEG) spectral exponent (i.e., exponential decay of power over frequency) has gained increasing attention as a novel marker of consciousness (Colombo et al. 2019; Lendner et al. 2020; Maschke, Duclos, Owen, et al. 2022; Colombo et al. 2023; Leroy et al. 2023). Especially in the absence of oscillatory peaks – which is a common phenomenon in disorders of consciousness (DOC) – our group has previously demonstrated diagnostic power of the spectral exponent above and beyond oscillatory power (Maschke, Duclos, Owen, et al. 2022). Besides the spectral exponent, spatial exponents – especially the anteriorization of alpha power – have been widely described as a marker for loss of consciousness in general anesthesia (Purdon et al. 2013; Vijayan et al. 2013; Purdon et al. 2015; Scheinin et al. 2018), sleep (De Gennaro et al. 2001) and DOC (Colombo et al. 2023).

Although Colombo et al. (2023) concluded that alpha power ‘cannot be a general marker of consciousness’, previous large-scale studies showed high diagnostic value of alpha power for patients in a DOC (Sitt et al. 2014; Engemann et al. 2018). There are three possible explanations for these contradictory findings. First, previously used large-scale datasets of DOC (Sitt et al. 2014; Engemann et al. 2018) contained patients with different etiologies which were not analyzed separately. The co-occurrence of lower levels of consciousness and suppressed broadband power in anoxic patients could induce a spurious diagnostic effect of alpha power when grouped with non-anoxic patients. The diagnostic effect of alpha power could therefore be driven by electrophysiological differences in etiologies, rather than etiology-independent levels of consciousness. Second, candidate markers of consciousness are widely validated against level of consciousness, as assessed using behavioral tools such as the CRS-R score (Kalmar and Giacino 2005). However, solely using behavioral tools for patient stratification is insufficient to detect capacity for consciousness despite unresponsiveness and could yield markers of consciousness which are more sensitive to behavioral responsiveness than the actual level of – or capacity for consciousness. Thus, alpha power could be an index of level of behavioral responsiveness, as shown by Sitt et al. (2014) and Engemann et al. (2018), rather than a reliable marker of capacity of consciousness (Colombo et al. 2023). Third, whereas alpha power is often defined as the relative contribution of the alpha bandwidth to the broadband power (herein called relative alpha power), Colombo et al. (2023) estimated power as the absolute amount of power in a given range (i.e., absolute alpha power). While relative alpha power normalizes for suppression in broadband activity, absolute alpha power is highly influenced by the amount of broadband background activity. Thus, different observations in the role of alpha power for the assessment of levels of consciousness could be driven by the different behavior of relative and absolute alpha power and their interaction with broadband background activity.

In addition to spatial and spectral properties, the temporal architecture of EEG signal has gained increasing attention for the evaluation of consciousness. Specifically, reduced spontaneous and evoked signal complexity marks unconsciousness resulting from anesthesia (Bruhn et al. 2000; Zhang et al. 2001; Jordan et al. 2008; Sarasso et al. 2015; Schartner et al. 2015), sleep (Burioka et al. 2005; Mateos et al. 2018), epilepsy (Mateos et al. 2018) and in disorders of consciousness (Sarà and Pistoia 2010; Gosseries et al. 2011; Casali et al. 2013; Sitt et al. 2014; Stefan et al. 2018; Lei et al. 2022). However, it remains unclear whether EEG complexity is an independent marker of level of consciousness, or epiphenomenal to changes in the spectral exponent. Although many studies have demonstrated a strong correlation between spectral exponent and spontaneous (Medel et al. 2020; Alnes et al. 2021; Maschke, Duclos, Owen, et al. 2022) and evoked signal complexity (Colombo et al. 2019; Maschke et al. 2023), most studies of signal complexity do not concurrently analyze changes in the spectral exponent.

In this study, we aim to shed light on the gap between those contradictory results by validating and contrasting the proposed markers of consciousness, specifically: 1) absolute alpha power; 2) relative alpha power; 3) the spectral exponent; and 4) the alpha posterior-anterior ratio (PAR); on a large-scale dataset of 260 patients in a DOC. Additionally, we calculate measures of complexity (i.e., Lempel-Ziv complexity), and assess their relationship to the four proposed markers of consciousness.

We compare the proposed markers against their ability to index level of and capacity for consciousness (i.e., diagnostic and prognostic value). Following Colombo et al. (2023), we hypothesize etiology-dependent differences in the diagnostic feature performance: while alpha power is expected to index levels of consciousness in anoxic patients, the spectral exponent and spatial gradients are hypothesized to generalize to non-anoxic patients and patients with other etiologies. In addition, we hypothesize that loss of signal complexity in lower levels of consciousness is epiphenomenal to changes in the spectral exponent, rather than an actual change of complexity. Alpha power is still widely used as a marker of level of consciousness in research, as well in clinical applications. This study has the potential to shed further light on the current debate about the identification and validation of neurophysiological markers of consciousness.

## 3 Materials and methods

### 3.1 Dataset

We analyzed an existing dataset containing resting-state EEG recordings from 303 patients with a DOC. All data was recorded in the Pitié - Salpêtrière hospital, Paris France. Consent was provided by the patients’ legal representative according to the Declaration of Helsinki. This study was approved by the ethical committee of the Pitié - Salpêtrière hospital under the code ‘Recherche en soins courants’. EEG data was acquired using a 256 channel GSN-HydroCel-257 sensor net.

### 3.2 Preprocessing

Data preprocessing was performed using MNE-python software (Gramfort et al. 2013). All data was downsampled to 250 Hz and filtered between 1 and 45 Hz. Notch filters were applied at 50 and 100 Hz. Electrodes which contained no signal or non-physiological artifacts were manually selected by a trained experimenter and removed from subsequent analysis. Due to spatial clustering of bad channels caused by small bandages or patches, removed channels were not interpolated. Data was average referenced and 62 non-brain channels were removed, yielding a maximum of 195 remaining channels. Data was epoched in 1 second windows without overlap. Bad epochs were dropped based on a maximal acceptable peak-to-peak amplitude of 200 µV. A maximum of 5 minutes preprocessed EEG was kept for subsequent analysis. From the initial 303 subjects, 33 subjects were rejected from subsequent analysis due to poor data quality and less than 2 minutes of clean data. Ten patients were excluded from the etiology-based analysis due to the co-occurrence of anoxic and non-anoxic brain injury (see Figure 1 for a graphic representation of participant inclusion). The remaining 260 patients (156 male, 102 female, 2 unknown, mean age 47.25 ± 17.25, 187 acute, 70 chronic) had an average of 185.84 ± 8.41 channels and 252.68 ± 48.99 seconds of data. No difference was observed in the number of epochs or channels between different etiologies or diagnostic groups.

**Figure 1.**
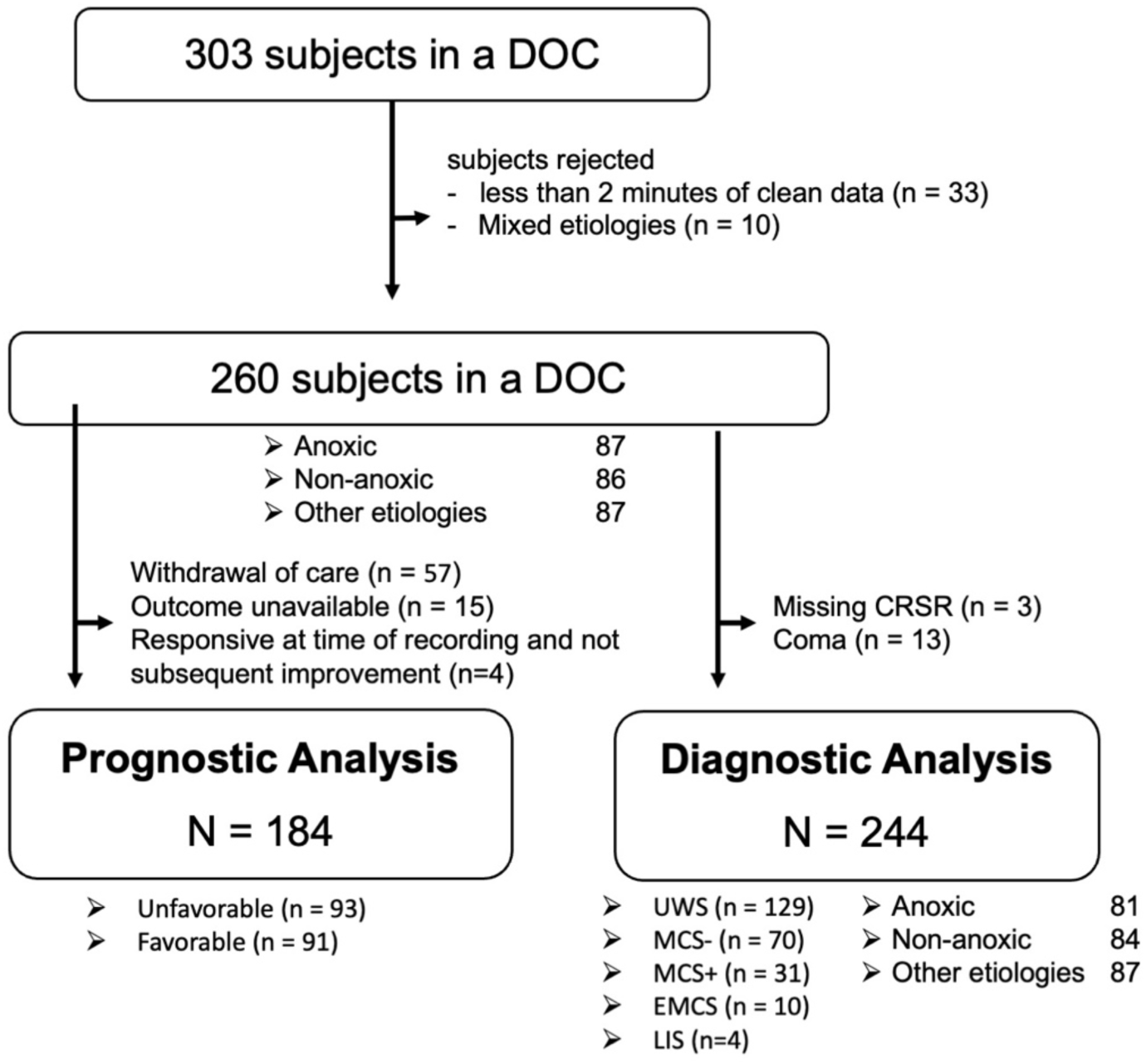
Visual representation of patients included in the diagnostic and prognostic analysis in this study. CRSR: Coma Recovery Scale Revised, UWS: Unresponsive Wakefulness Syndrome, MCS: Minimally Conscious State, EMCS: Emergence, LIS: Locked in syndrome.

### 3.3 Patient Etiology

This study aims to compare the diagnostic value of candidate markers of consciousness in different etiologies, especially anoxic and non-anoxic injuries. We therefore split the patients in three groups, according to their etiology. From the 260 patients included in this study, 87 patients suffered from an anoxic brain injury, 86 patients suffered a non-anoxic injury (n= 63 TBI and n=23 stroke). 86 patients were unresponsive following a variety of other etiologies including encephalitis, encephalomyelitis, leukoencephalopathy, hypoglycemia, post-operative complications, intoxication, toxoplasmosis, meningioma, hyperthermia, malaria, undefined etiology, unconsciousness following COVID-19, Guillain Barré and septic chocs.

### 3.4 Patient Diagnosis

Patients’ level of consciousness was assessed on the day of recording by a trained experimenter using the CRS-R score (Kalmar and Giacino 2005). At the time of EEG data acquisition, 13 patients were in a coma, 129 patients were diagnosed with UWS, 70 patients were in MCS-, 31 patients were in MCS+, and 10 patients were in EMCS (see Table 1 for demographic information on each group). Four patients were unresponsive following from Guillain Barré and were therefore classified to be in a locked in syndrome. Three patients had no recorded diagnosis and were excluded from the diagnostic analysis. As we cannot exclude the presence of consciousness despite complete unresponsiveness, the 13 patients in a coma were excluded from the diagnostic analysis (see Figure 1 for a graphic representation of participant inclusion).

**Table 1:**
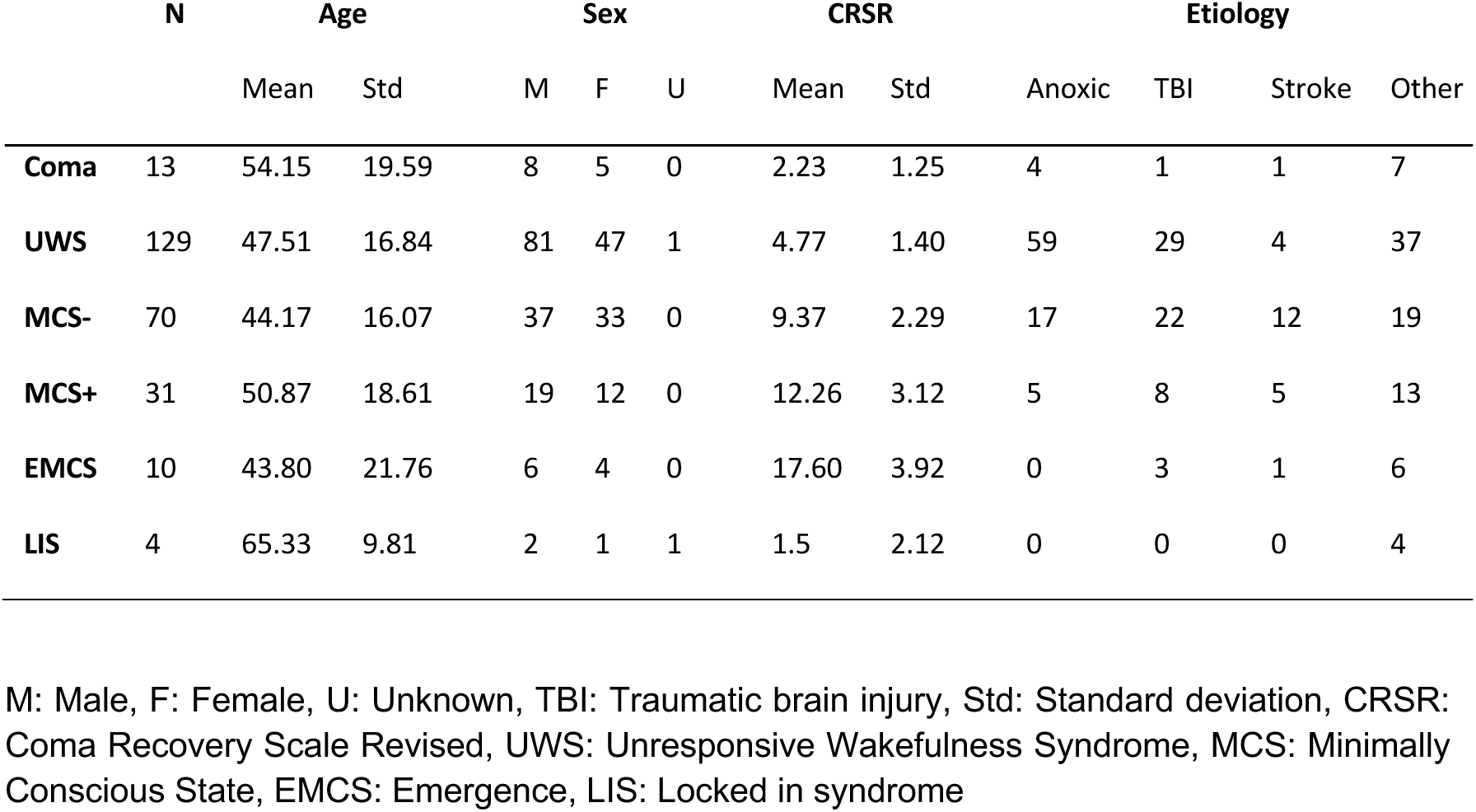
Demographic information for diagnostic analysis.

|  | N | Age |  | Sex |  |  | CRSR |  | Etiology |  |  |  |
| --- | --- | --- | --- | --- | --- | --- | --- | --- | --- | --- | --- | --- |
|  |  | Mean | Std | M | F | U | Mean | Std | Anoxic | TBI | Stroke | Other |
| **Coma** | 13 | 54.15 | 19.59 | 8 | 5 | 0 | 2.23 | 1.25 | 4 | 1 | 1 | 7 |
| **UWS** | 129 | 47.51 | 16.84 | 81 | 47 | 1 | 4.77 | 1.40 | 59 | 29 | 4 | 37 |
| **MCS-** | 70 | 44.17 | 16.07 | 37 | 33 | 0 | 9.37 | 2.29 | 17 | 22 | 12 | 19 |
| **MCS+** | 31 | 50.87 | 18.61 | 19 | 12 | 0 | 12.26 | 3.12 | 5 | 8 | 5 | 13 |
| **EMCS** | 10 | 43.80 | 21.76 | 6 | 4 | 0 | 17.60 | 3.92 | 0 | 3 | 1 | 6 |
| **LIS** | 4 | 65.33 | 9.81 | 2 | 1 | 1 | 1.5 | 2.12 | 0 | 0 | 0 | 4 |

M: Male, F: Female, U: Unknown, TBI: Traumatic brain injury, Std: Standard deviation, CRSR: Coma Recovery Scale Revised, UWS: Unresponsive Wakefulness Syndrome, MCS: Minimally Conscious State, EMCS: Emergence, LIS: Locked in syndrome

### 3.5 Patient Prognosis

Each patient’s level of recovery was evaluated 6- and 12-months following injury using a pseudo CRS-R via phone call (Kalmar and Giacino 2005) (n= 245 at 6 months, n= 217 at 12 months) and the GOS-E (Jennett and Bond 1975) (n= 253 at 6 months, n= 213 at 12 months). From the initial 260 patients, 57 patients died due to withdrawal of life supporting treatment. Due to the inability to assess those patients’ ‘natural’ progression post-injury and to avoid confounding factors such as patient’s and caregivers’ socioeconomic status, those patients were excluded from the prognostic analysis (see Figure 1). For 15 patients, the outcome measure was not available yielding 188 patients with known functional outcome.

For the purpose of this study, functional outcome was defined as the maximal level of responsiveness reached within one year post injury (i.e., patients who regained responsiveness, but later deceased were labeled as ’recovered responsiveness’, as the reason of death might not be injury-related). As command following is the criteria separating MCS-from MCS+ (Kalmar and Giacino 2005), patients in an MCS+ and patients who regained consciousness were grouped into the prognostic category ’recovered responsiveness’ (i.e., Positive Outcome). Patients who did not regain responsiveness (i.e., remained in MCS-, UWS or Coma) and patients who died (not caused by withdrawal of life sustaining treatment) were grouped into the prognostic category named ‘did not recover responsiveness’ (i.e., Negative Outcome). Patients who were already in an MCS+ or EMCS at time of recording and did not improve over one year post injury were excluded from the prognostic analysis (n=4).

Using the GOS-E score, a positive outcome corresponds to a score above 2, while a negative outcome corresponds to values of 1 and 2 (see supplementary Figure S1 for the comparison of prognosis using CRS-R and GOS-E score). For one patient, the outcome was assessed based on only one score, due to missing values in the second test (see supplementary Figure S1).

A total of 184 patients were included in the prognostic analysis (see Figure 1), from which 91 recovered responsiveness and 93 did not recover responsiveness (see Table 2 for prognostic information on each group, see supplementary Figure S2 for the prognostic groups split by original diagnosis, medical state and patient’s sex). No significant diagnostic and prognostic group-difference was found in patient’s age and sex.

**Table 2:** Demographic information for prognostic analysis, spitted by prognostic groups.

| Outcome | N | Age |  | Sex |  |  | Etiology |  |  |  |
| --- | --- | --- | --- | --- | --- | --- | --- | --- | --- | --- |
|  |  | Mean | Std | M | F | U | Anoxic | TBI | Stroke | Other |
| Positive | 91 | 44.50 | 17.47 | 53 | 38 | 0 | 18 | 31 | 11 | 31 |
| Negative | 93 | 48.68 | 17.21 | 58 | 35 | 0 | 35 | 22 | 9 | 27 |

Positive: Patients who recovered responsiveness within one year post recording. Negative: Patients who did not recover responsiveness within one year post recording. M: Male, F: Female, U: Unknown, TBI: Traumatic brain injury, Std: Standard deviation

### 3.6 Feature extraction

#### 3.6.1 Spectral features

##### Spectral power

Power spectral density was calculated on each epoch and channel individually using the Welch method implementation from MNE python (Gramfort et al. 2013) with a 1 second windows and 50% overlap (*fmin=1, fmax=45, n_fft=250, n_overlap=125*). Frequency bands were defined as delta (1-4 Hz), theta (4-8Hz), alpha (8-13Hz), low beta (13-20Hz) and high beta (20-30 Hz). Prior to power calculation, power spectral density was averaged over epochs, yielding one power spectrum per channel. The power analysis (i.e., absolute, relative and flat power) was performed on each channel individually and subsequently averaged over space.

Absolute power in each frequency band was defined as log10 of the area under the PSD curve in the respective frequency band. Relative power was defined as the area under the PSD curve in the respective frequency band, divided by the area of the full spectrum (1-45 Hz). Relative power thus reflects a percentual contribution of a specific frequency range to the whole spectrum. In contrast to absolute alpha power, which is affected by global power suppression, relative contribution of alpha power to the power spectrum corrects for differences in broadband power and captures differences in the shape of the power-spectral density, rather than the absolute height of a specific band. Flattened power was calculated using the ‘fitting oscillations and one over f’ fooof._spectrum_flat function (Donoghue et al. 2020). This function subtracts the aperiodic component from the power spectrum, leaving only the oscillatory peaks. Flattened power was defined as the are under the flattened power spectral density in the respective frequency band.

##### Posterior-Anterior Ratio

Posterior-Anterior ratio (PAR) was defined as the geometric mean of all posterior values of absolute power (i.e., area under the PSD curve without log) divided by the geometric mean of anterior values, following Colombo et al. (2023). While anterior-dominated activity results in a PAR between 0 and 1, with 0 being total anteriorization, values above 1 indicate posterior-dominant activity. The posterior-anterior regions were split along the line of Cz, electrodes on the midline were not included in the analysis (see supplementary Figure S3). The Python function for the calculation of the posterior-anterior ratio is available at https://github.com/BIAPT/Markers.

##### Aperiodic signal components

The aperiodic signal components (i.e., spectral exponent and offset) were calculated using the fixed mode of the ‘fitting oscillations and one over f’ algorithm (Donoghue et al. 2020). To avoid overfitting and detection of spurious oscillation, hyperparameters for the peak detection were used as recommended by Gerster et al. (2022) (*min_peak_height=0.1, max_n_peaks=3, peak_width_limits=(2, 5.0)*). Aperiodic components were calculated on the power spectrum of each channel individually and averaged subsequently over space.

##### Complexity

Signal complexity was calculated using Lempel-Ziv complexity (Lempel and Ziv 1976). EEG signal was epoched in 10 second non-overlapping windows and was discretized using the signal mean as a threshold. Signal complexity was calculated for every channel and epoch individually and averaged over epochs subsequently. To account for possible influence of the signals’ spectral properties, LZC was estimated using two different methods for normalization: 1) shuffle normalization and 2) phase normalization.

In shuffle normalized LZC, the complexity of the binarized EEG timeseries was normalized using the mean after randomly shuffling the previously obtained binary series 100 times. While complexity of 0 indicate a repetitive and easily compressible signal, complex signals which not easily compressible approach complexity values of 1.

For the phase normalized LZC, each signals’ complexity was normalized using the mean complexity of surrogate signals. Phase-randomization has previously been used as a tool to decorrelate changes in complexity from pure spectral signal changes (Schartner et al. 2017; Toker et al. 2022). The surrogate signal was obtained by randomizing the phase of the original time series while maintaining most of its spectral properties. 100 surrogate signals were obtained for every epoch and channel individually. Complexity values above 1 indicate a higher signal complexity, relative to other signals with comparable spectral properties. Values below one indicates a relatively lower signal complexity. Our custom python code for the calculation of shuffle- and phase-normalized LZC is available at https://github.com/BIAPT/Markers.

### 3.7 Diagnostic and prognostic analysis

To assess each features’ ability to index levels of consciousness we used five different approaches, summarized in Figure 2: 1) the correlation with patients’ diagnosed level of consciousness (UWS, MCS-, MCS+, EMCS); 2) the correlation with patient’s total CRS-R score; 3) the distinction between patients in a UWS and patients with higher levels of consciousness (MCS-, MCS+ and EMCS); 4) the distinction between unresponsive patients (UWS and MCS-) and patients with some level of responsiveness (MCS + and EMCS); and 5) the distinction patients according to their outcome (see Figure 2). To account for the possible influence of etiology, all analyses were performed first on the totality of patients and then in subgroups according to patient’s etiology (i.e., anoxic, non-anoxic and other injuries).

**Figure 2:**
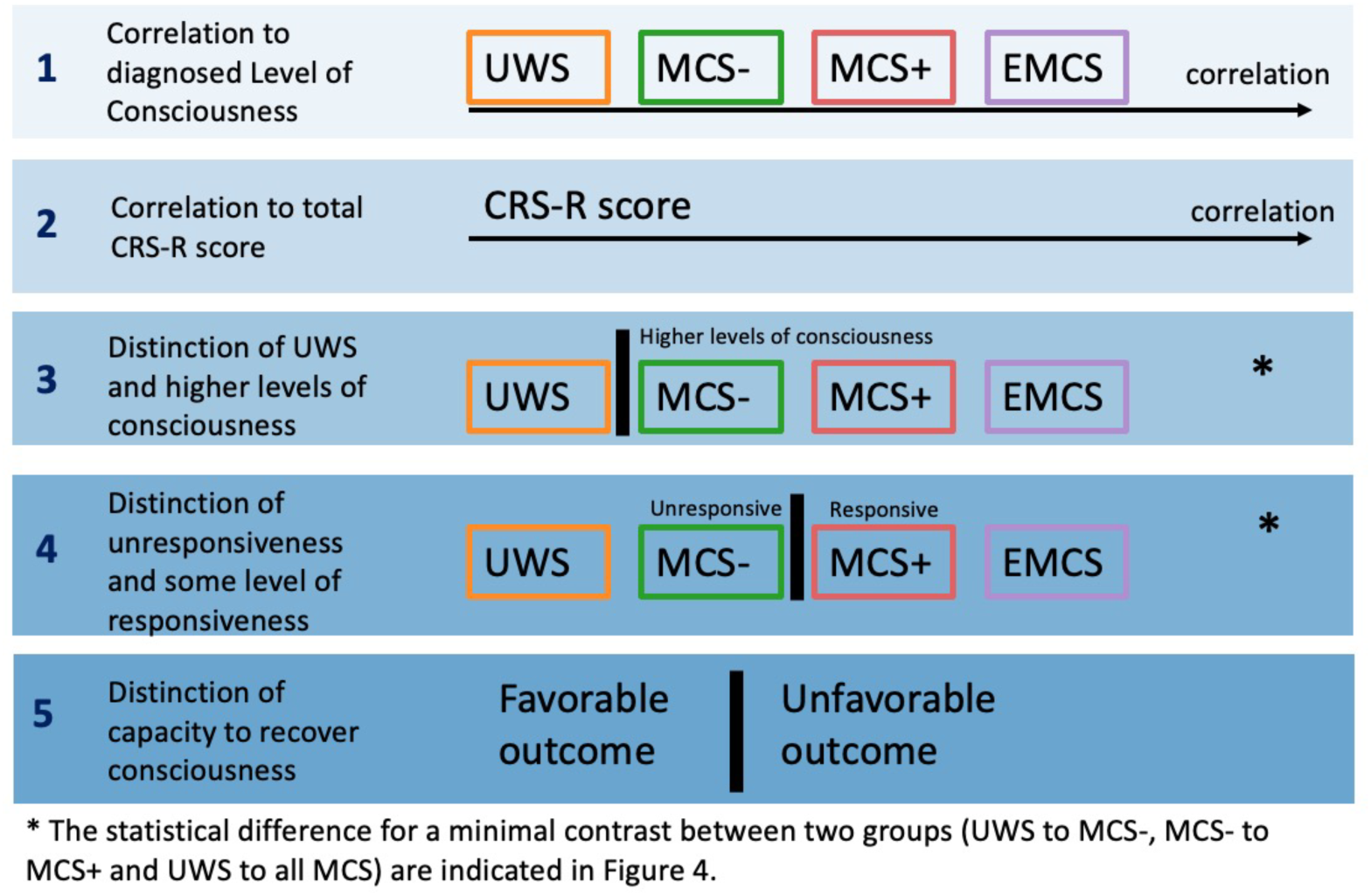
Overview of analysis used to assess the value of candidate features for the assessment of level of (analysis 1-4) and capacity for consciousness (analysis 5). CRSR: Coma Recovery Scale Revised, UWS: Unresponsive Wakefulness Syndrome, MCS: Minimally Conscious State, EMCS: Emergence

In the above-described analysis, patients with different diagnoses were grouped together as ‘higher levels of consciousness’ (i.e., MCS-, MCS+ and EMCS) or ‘some level of responsiveness (i.e., MCS+ and EMCS). Although this approach provides a larger contrast, it also introduces a variety of confounding factors, as patients do not differ solely in their level of consciousness. Within the category of MCS alone, patients may range from completely unconsciousness despite residual cortical activity and overt behavior to self-conscious individuals who lack the executive capacity to respond (Naccache 2018). To maximally reduce confounding factors, we additionally investigate the minimal contrast between adjacent diagnostic groups (i.e., UWS to MCS-, UWS to all MCS, MCS-to MCS+).

### 3.8 Statistical analysis

Correlation with the patient’s level of consciousness and total CRS-R score was performed using a partial Spearman rank partial correlation (Spearman’s Rho), implemented by the pingouin package (Vallat 2018). Features such as the spectral exponent (Voytek et al. 2015) and alpha PAR (Chiang et al. 2011) are variable over lifespan. To account for interactions between candidate measures and patient age, all correlations were performed using age as a covariate. A feature’s ability to differentiate patients, according to analysis 3-5 was assessed using a Mann-Whitney-U-test. We corrected for multiple comparisons using the group-wise Holm correction (i.e., correction was performed for all seven metrics of interest within one etiology). In the first analysis, spearman correlation was applied for the relative rank of patient diagnosis (0: UWS, 1: MCS-, 2: MCS+ and 3: EMCS). In the second analysis, we performed the Spearman rank test using patients’ total CRS-R score.

## 4 Results

We first report the effect of grouping different etiologies of unresponsive patients on the candidate EEG markers. Subsequently, we contrast the diagnostic value of alpha power between its various instantiations (i.e., absolute, relative and flattened alpha power). We then compare the diagnostic value of spatial and spectral gradients (i.e., alpha PAR and the spectral exponent). Finally, we explore the influence of spectral properties on signal complexity. A summary of results from all five conducted analysis is presented in Figure 3, the corresponding statistical tables can be found in the supplementary material (see supplementary Table S1-S5).

**Figure 3:**
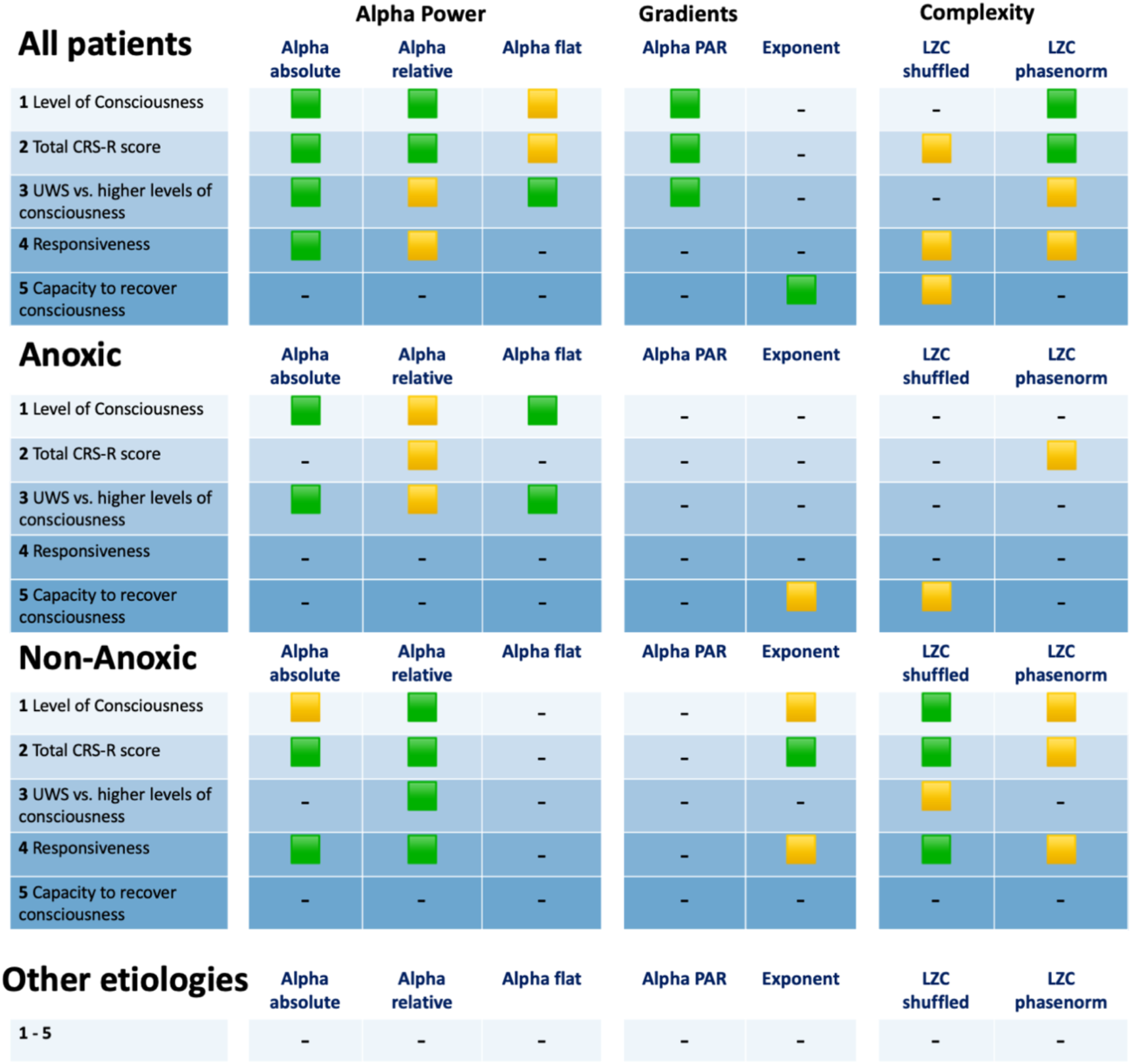
**Overview of all analysis results** first presented for all etiologies, then split into groups based on patient etiology. Green squares represent significant effects. Yellow squares indicate effects which lost significance after correcting for multiple comparison. All statistical tables can be found in the supplementary Tables S1-S5. PAR: Posterior anterior ratio, LZC Lempel-Ziv Complexity.

### 4.1 Grouping unresponsive patients from different etiologies can induce spurious correlations or obscure existing markers of diagnosis

Figure 3 illustrates a discrepancy between effects observed on the whole-group level and diagnostic value of candidate EEG markers on individual etiologies. We present two examples of diagnostic effects which were observed on the whole group of unresponsive patients but did not generalize to individual subgroups.

When analyzing patients from all etiologies grouped together, absolute alpha power significantly correlated with patient’s level of consciousness (r(238) = 0.31, p < 0.001) (analysis 1). However, after splitting patients according to their etiology, the correlation between the levels of consciousness and absolute alpha power remained only for anoxic patients (r(79) = 0.38, p < 0.01) (analysis 1). For non-anoxic patients, the relation between absolute alpha power and levels of consciousness strongly weakened and lost significance after p-value correction. No significant correlation was found for patients with other etiologies. A post-hoc analysis confirmed that anoxic patients in this dataset were characterized by a significantly lower CRS-R score (U = 2235, p<0.001) and broadband power (U = 2291, p<0.001), compared to non-anoxic patients. The lower broadband power resulted also in significantly reduced absolute alpha power (U = 2491, p<0.001) in anoxic patients. Our results demonstrate that lower CRS-R score and reduced broadband power in anoxic patients can induce a spurious group-level diagnostic value of an EEG marker for patients in a DOC, which might vanish when considering etiologies individually.

An even stronger effect was observed for alpha PAR, which had diagnostic value solely on a whole-group level (i.e., analysis 1-3). Similar to alpha power, a post-hoc analysis confirmed that anoxic patients were characterized by a significantly more posterior-dominant alpha activity (i.e., lower alpha PAR) (U = 2285, p<0.001), compared to non-anoxic patients. When performing the analysis on individual etiologies independently, the diagnostic effects vanished for all groups. Beyond inducing spurious correlations, grouping all unresponsive subjects together also overshadowed existing etiology-specific diagnostic markers. When analyzing all etiologies together, the spectral exponent did not have any diagnostic value (analysis 1-3). However, splitting subjects by etiology revealed a strong link between the spectral exponent and non-anoxic patients’ CRS-R score (r(82) = 0.33, p<0.05) (analysis 2). In line with our previous findings (Maschke, Duclos, Owen, et al. 2022), a flatter spectral exponent was indicative for higher levels of consciousness.

As recommended by Colombo et al. (2023), patients were split in anoxic and non-anoxic (i.e., stroke and TBI) patients. The correlation with patient’s level of consciousness was reproduced across patients who suffered a traumatic brain injury and stroke patients. Results are provided in the supplementary material (see supplementary Figure S4, see Discussion).

### 4.2 Alpha power has a higher diagnostic importance for anoxic compared to non-anoxic patients

When splitting the patients according to their etiology into anoxic (n = 81), non-anoxic (n = 84) and patients with other etiologies (n = 79), absolute alpha power maintained strongest diagnostic value for anoxic patients. Absolute alpha power correlated to the level of consciousness of anoxic patients (r(79) = 0.38, p<0.01) (analysis 1) and significantly distinguished UWS patients from patients with higher levels of consciousness (U = 321, p < 0.01) (analysis 3) (see Figure 4 and 5). In line with previous research (Colombo et al. 2023) higher levels of alpha power indicated higher levels of consciousness for anoxic patients. In addition, absolute alpha power significantly differentiated anoxic patients in a UWS from MCS-(U = 233, p < 0.01) and patients in a UWS from all MCS patients (U = 321, p < 0.001).

**Figure 4.**
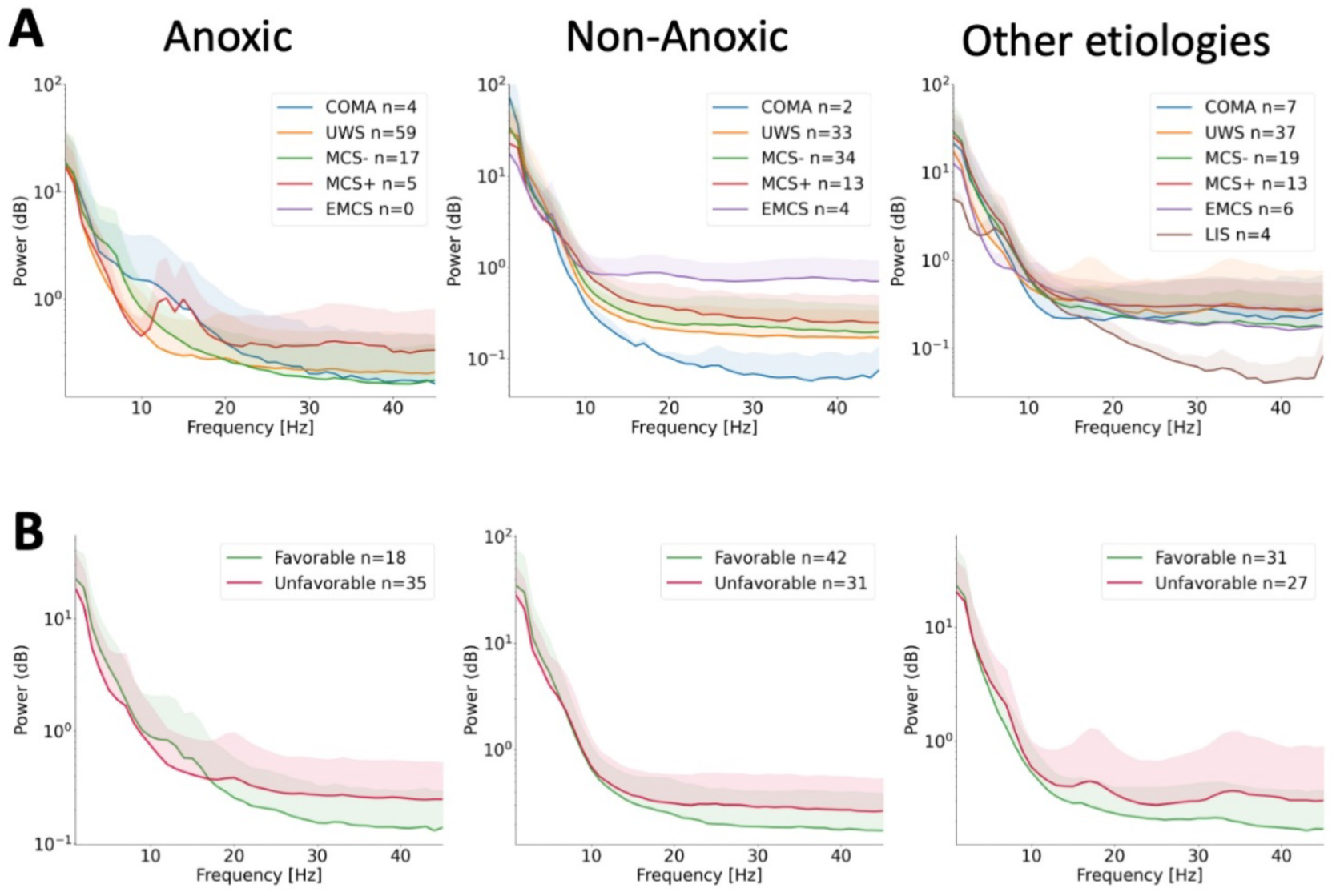
Spectrograms for different etiologies and diagnostic and prognostic groups. Power spectral density and of anoxic (left) and non-anoxic (middle) and (right) patients who were unresponsive due to other etiologies. Solid lines represent the mean of the (A) diagnostic and (B) prognostic groups. Shaded areas are the one-sided standard deviation above the mean. UWS: Unresponsive Wakefulness Syndrome, MCS: Minimally Conscious State, EMCS: Emergence, LIS: Locked in syndrome. Coma patients were not included in the diagnostic analysis and are presented for purpose of visualization only (see Methods)

**Figure 5.**
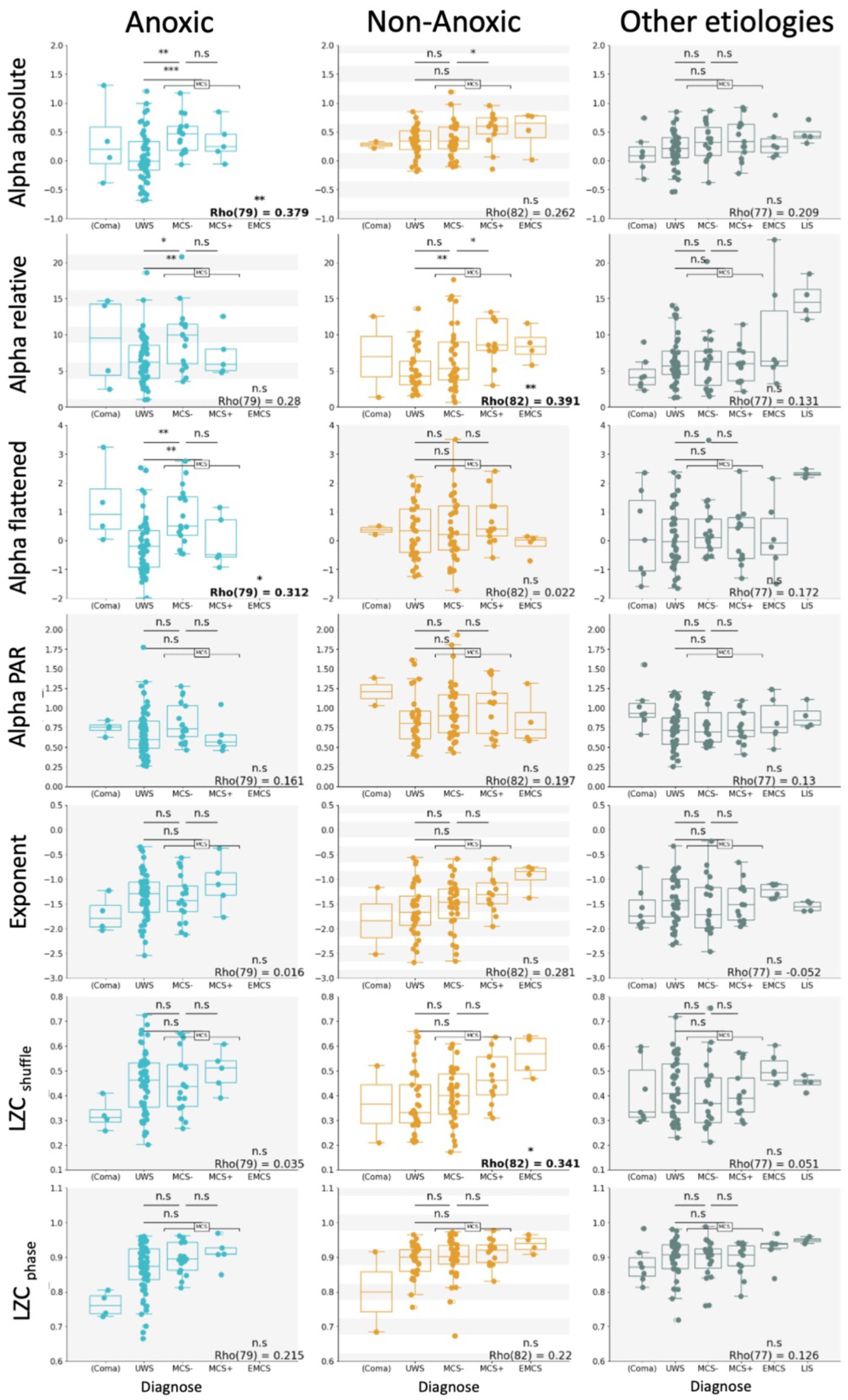
Correlation between spectral properties and patients’ levels of consciousness. Correlation between patients’ diagnosis, and all candidate EEG features. Spearman rank correlation value and degrees of freedom are indicated for each correlation. Each dot corresponds to an individual patient. Corrected significance values are indicated above, with * indicating p< 0.05, ** being p < 0.01, *** being p< 0.001 and n.s. indicating no significance. All non-significant correlations are shaded with grey background. A background with grey and white lines indicates loss of significance after p-value correction. A post-hoc test was performed to assess the minimal contrast between diagnostic groups. P-values are indicated above the bars to differentiate UWS from MCS-, MCS-from MCS+ (first row) and UWS from all MCS (second row). Coma patients were not included in the diagnostic analysis and are presented for purpose of visualization only (see Methods). UWS: Unresponsive Wakefulness Syndrome, MCS: Minimally Conscious State, EMCS: Emergence, LIS: Locked in syndrome, PAR: Posterior anterior ratio, LZC Lempel-Ziv Complexity.

For non-anoxic patients, the correlation between absolute alpha power and patient’s level of consciousness (analysis 1) lost significance after p-value correction. However, a significant correlation remained between absolute alpha power and non-anoxic patients’ CRS-R score (r(82) = 0.31, p < 0.05) (analysis 2). In addition, absolute alpha power differentiated non-anoxic patients according to their level of responsiveness (U = 320, p < 0.05) (analysis 4). There was no significant difference in absolute alpha power between non-anoxic patients in a UWS and MCS- or UWS and MCS. However, compared to patients in an MCS-absolute alpha power was significantly increased for non-anoxic patients in a MCS+ (U = 131, p < 0.05). No diagnostic effects were present for patients with other etiologies. Alpha power had no prognostic value for any of the etiology groups.

Compared to absolute alpha power, relative alpha power clearly showed the strongest diagnostic value for non-anoxic patients, with significant effects in analysis 1 to 4. Relative alpha power was significantly correlated with non-anoxic patient’s level of consciousness (r(82) = 0.39, p<0.01) (analysis 1) and CRS-R score (r(82) = 0.44, p < 0.001) (analysis 2) (see Figure 4 and 5). In addition, relative alpha power distinguished non-anoxic UWS patients from patients with higher levels of consciousness (U = 522, p < 0.05) (analysis 3) and stratified patients according to their level of responsiveness (U = 269, p < 0.01) (analysis 4). In line with this, relative alpha power significantly differentiated non-anoxic patients in a UWS from MCS (U = 500, p<0.01) and patients in a MCS-from MCS+ (U = 128, p<0.05). In the group of anoxic subjects, all diagnostic effects from analysis 1-4 lost significance after p-value correction. Only the minimal contrast comparison between UWS and MCS-(U = 267, p < 0.05) and UWS and all MCS (U = 396, p<0.01) showed significance (see Figure 5). A visualization of spatial distribution of absolute and relative alpha power in UWS and MCS is provided in the supplementary material (see supplementary Figure S5).

Flattening alpha power disentangled the effect of alpha oscillation from the background activity or spectral exponent of the power spectral density. After removing the background activity from alpha power (herein called ‘flattened alpha power’), this feature exhibited diagnostic value only in anoxic patients. Flattened alpha power correlated with anoxic patients’ level of consciousness (r(79) = 0.31, p < 0.05) (analysis 1) and was able to significantly differentiate patients in a UWS from patients with higher levels of consciousness (U = 363, p < 0.05) (analysis 3) (see Figure 5). In addition, flattened alpha power significantly distinguished anoxic patients in an UWS from MCS-(U=231, p<0.01) and UWS from all MCS (U=363, p<0.01) (see Figure 5). No diagnostic effect was found for non-anoxic patients and patients with other etiologies.

In summary, absolute alpha power had diagnostic value for both anoxic and non-anoxic subjects. While anoxic patients showed effects in the differentiation of levels of consciousness, the effect in non-anoxic patients was more expressed in differentiating responsiveness. Meanwhile, relative alpha power showed diagnostic effects only for non-anoxic patients. Separating alpha power from the spectral exponent limited the diagnostic value to anoxic patients. In a post-hoc analysis we looked at the relative power in all frequency bands. In line with previous research (Sitt et al. 2014; Engemann et al. 2018) we showed that patient’s diagnosis correlated negatively with relative delta power and positively with all relative power bands above 8 Hz (see supplementary Figure S6). This indicates that diagnostic value of relative power in non-anoxic patients is largely attributable to the spectral exponent, rather than the alpha peak. Our results confirm the hypothesis that alpha power has higher diagnostic value for anoxic patients, compared to non-anoxic patients.

### 4.3 The EEG spectral exponent has more diagnostic value than the spatial gradient

When analyzing patients from all etiologies grouped together, the alpha PAR showed high diagnostic value, being significantly correlated with patient’s level of consciousness (r(238) = 0.19, p < 0.05) (analysis 1) and CRS-R score (r(238) = 0.19, p < 0.05) (analysis 2). In addition, alpha PAR significantly differentiated patients in a UWS from patients with higher levels of consciousness (U = 5404, p < 0.01) (analysis 3). In line with Colombo et al (2023), lower CRS-R scores aligned with stronger alpha anteriorization. However, after splitting patients according to their etiology, all effects vanished for all patient groups (see Figure 5).

In contrast to the spatial gradient, the spectral exponent did not show diagnostic value on a whole-group level. However, the spectral exponent significantly correlated with non-anoxic patients’ CRS-R score (r(82)=0.33, p<0.05) (analysis 2). In line with previous research (Maschke, Duclos, Owen, et al. 2022), a flatter exponent indicated a higher level of consciousness (see Figure 4 and 5). The correlation between the spectral exponent and non-anoxic patients’ level of consciousness as well as its ability to distinguish responsive from unresponsive patients lost significance after p-value correction (see Figure 3 and 5).

In addition, it is interesting to note the potential prognostic value of the spectral exponent for anoxic patients. Even though this difference did not remain significant after p-value correction, patients with an unfavorable outcome were characterized by a significantly flatter spectral exponent compared to patients with a favorable outcome (U = 175, p uncorrected < 0.01, p corrected = 0.06). In addition, a post-hoc analysis revealed a positive correlation with anoxic patients’ maximal GOS-E score (r(51) = -0.30, p uncorrected < 0.05). Despite a strong correlation between the EEG spectral exponent and offset (r(259) = -0.81, p<0.001), the spectral exponent showed consistently higher diagnostic value than the offset.

### 4.4 Changes in signal complexity can be largely attributed to changes in the spectral exponent

In line with previous research pointing out the strong relationship between the spectral exponent and Lempel-Ziv Complexity (Medel et al. 2020), the spectral exponent showed a high correlation with the shuffle-normalized LZC (r(259) = 0.93, p<0.001). Normalizing the signal with the phase shuffled surrogates reduced this relationship (r(269) = 0.52, p<0.001), yielding measures of complexity which were more independent of spectral properties.

Similar to the results from the spectral exponent, shuffle-normalized LZC did correlate with non-anoxic patients’ diagnosed level of consciousness (r(82) = 0.34, p<0.05) (analysis 1) and CRS-R score (r(82) = 0.39, p < 0.01) (analysis 2). It further significantly differentiated non-anoxic patients according to their responsiveness (U = 296, p < 0.05) (analysis 4). Correcting the Lempel-Ziv complexity for phase differences weakened all previously significant diagnostic value for non-anoxic patients. Thereby, all diagnostic effects of phase-shuffled LZC lost significance after p-value correction. Likewise, correcting the correlation between shuffle-normalized LZC and patients’ level of consciousness and CRS-R score by the spectral exponent (i.e., using partial correlation) made the diagnostic effect vanish.

Taken together, whereas shuffle-normalized LZC reproduced similar effects as seen in the spectral exponent (see Figure 5), phase-normalization (i.e., reducing the influence of spectral properties on the complexity) reduced the effects or eliminated the observed diagnostic value. However, shuffle-normalized LZC exhibited a generally stronger diagnostic effect compared to the spectral slope (analysis 1-4). Our results indicate that the diagnostic effect of LZC can be largely, but not exclusively, attributed to spectral changes – specifically changes in the spectral exponent – with flatter exponents indicating higher levels of complexity.

### 4.5 None of the candidate markers generalized to patients with other etiologies

None of the investigated markers had diagnostic or prognostic value that generalized to patients who were unresponsive due to other injuries (see Discussion).

## 5 Discussion

In this study, we investigated the diagnostic and prognostic value of three classes of EEG features, namely: 1) spectral power; 2) spectral and spatial gradients; and 3) signal complexity; for the assessment of unresponsive patients. Our results indicate that the diagnostic value of absolute and flat alpha power is most strongly expressed in anoxic patients, contributing to growing evidence that alpha power indexes the level of cortical suppression rather than level of consciousness. In contrast, changes in relative power had strong diagnostic effects for non-anoxic patients but were largely attributable to changes in the spectral exponent. The EEG spectral exponent had diagnostic value for non-anoxic patients only. Although the diagnostic value of signal complexity was largely attributable to spectral changes, some diagnostic value remained after correcting for spectral changes. Diagnostic value of signal complexity was largely, but not fully attributable to spectral changes, indicating a need to identify of measures of complexity which are more independent of spectral signal properties. None of the candidate measures generalized to patients with other etiologies.

### Etiology is a strong confounding factor in EEG markers of consciousness

Grouping unresponsive patients from different etiologies together can both induce spurious correlations and obscure existing etiology-based markers of diagnosis. When analyzing all unresponsive patients together, many of the candidate features including absolute alpha power, the alpha PAR and signal complexity showed diagnostic value for unresponsive patients. However, splitting patients into groups based on patients’ etiology eliminated the diagnostic value of these markers for individual etiologies. Compared to non-anoxic patients, anoxic patients are often characterized by an overall lower CRS-R score and reduced broadband activity (Estraneo et al. 2016; Snider et al. 2022; Colombo et al. 2023). Analyzing anoxic and non-anoxic patients together can therefore induce a spurious correlation driven solely by differences between etiologies, rather than level of consciousness. We confirm our hypothesis that putative EEG markers of consciousness have etiology-dependent differences in their association with patient diagnosis and recommend considering etiologies independently when searching for markers of level of and capacity for consciousness.

### Alpha power has higher diagnostic value for anoxic patients

Absolute alpha power was related to diagnosis in anoxic and non-anoxic patients. Absolute alpha power was more associated with levels of consciousness in anoxic patients and separated non-anoxic patients only on their level of responsiveness. In contrast, relative alpha power had high diagnostic value for non-anoxic, but not for anoxic patients. However, correcting alpha power for the background activity (i.e., subtracting the aperiodic component from the power spectral density) limited the diagnostic association to anoxic patients only. Thus, non-anoxic patients’ diagnostic value of relative alpha power and relative power in other frequency bands (see supplementary material Figure S6) was largely attributable to changes in the spectral exponent. We therefore confirm our hypothesis that alpha power has higher diagnostic value for anoxic patients and highlight the importance of considering the spectral exponent in power analyses. Our results support findings by Colombo et al (2023), who attributed loss of alpha power to global suppression of cortical activity in anoxic patients, rather than level of consciousness.

Previous studies (Sitt et al. 2014; Engemann et al. 2018; O’Donnell et al. 2021) demonstrated the diagnostic value of relative alpha power. The authors of Colombo et al. (2023) speculated that those findings could be attributed to the overrepresentation of anoxic patients and the use of neurobehavioral tools for patient stratification. However, the use of relative alpha power in those papers avoids a bias originated by the broadband power suppression of anoxic patients. Instead, our results indicate that the previously identified diagnostic and prognostic value of relative alpha power –specifically for non-anoxic patients– (Sitt et al. 2014; Engemann et al. 2018; O’Donnell et al. 2021) could be partially attributed to changes in the spectral exponent of their EEG.

### The EEG spectral exponent has diagnostic value for non-anoxic patients

Even though the EEG spectral exponent has been historically neglected, mounting evidence attributes changes in the spectral exponent as an index of altered states of consciousness (Colombo et al. 2019; Lendner et al. 2020; Colombo et al. 2023). In addition, our group previously demonstrated that patients in a DOC are often characterized by a total absence of oscillatory peaks (Maschke, Duclos, Owen, et al. 2022). In the current study, we validate the previously shown diagnostic value of the aperiodic component for non-anoxic patients (Maschke, Duclos, Owen, et al. 2022; Colombo et al. 2023). As demonstrated previously (Maschke, Duclos, Owen, et al. 2022; Colombo et al. 2023), higher levels of consciousness were indexed by a flatter spectral exponent. In line with previous results (Colombo et al. 2023), the spectral exponent had no diagnostic value for anoxic patients. None of the tested measures generalized to patients who did not suffer brain injury. In summary, the spectral exponent has diagnostic value for non-anoxic patients but does not generalize to anoxic subjects and patients with other etiologies.

### Alpha posterior-anterior ratio was not related to diagnosed level of consciousness

In contrast to the spectral exponent, spatial gradients (i.e. the alpha PAR) showed only little diagnostic value for non-anoxic patients. The positive correlation between the alpha PAR and non-anoxic patient’s total CRS-R score lost significance after p-value correction. Colombo et al. (2023) showed that alpha PAR had a stronger capacity to stratify levels of consciousness, compared to alpha power. In their study, alpha PAR indexed levels of consciousness in non-anoxic patients and the reference dataset of anesthetic-induced unconsciousness. In our study, we were not able to replicate the diagnostic value of alpha PAR. We discuss two possible explanations for the contradicting results.

First, the study by Colombo et al. (2023) quantified patients’ level of consciousness using a neuro-behavioral assessment, combining the CRS-R score (Giacino et al. 2004) (i.e., a behavioral assessment) with the perturbational complexity index (Casali et al. 2013) (i.e., a neurological assessment). While this allowed them to identify unresponsive patients with higher levels of consciousness, our study relied on behavioral assessment only. The reliance on behavioral proxies of consciousness likely underestimated levels of consciousness in our patients, leading to inability to detect diagnostic effect in alpha PAR. However, while the above argument concerns the evaluation of diagnostic value in all features (see Limitations), the inability to replicate results presented by Colombo et al. (2023) only concerned the alpha PAR. We thus consider a second explanation for the contradicting findings.

Alpha power is sensitive to demographic characteristics such as age and sex (Jaušovec and Jaušovec 2010; Chiang et al. 2011; Cave and Barry 2021). More specifically, woman have higher broadband power and stronger alpha activity – particularly in parietal and occipital regions (Jaušovec and Jaušovec 2010; Cave and Barry 2021). In addition, the alpha peak has a strong relation to age, moving towards more frontal regions across the lifespan (Chiang et al. 2011). Indeed, a post-hoc analysis on our data confirmed a significant correlation between age and the alpha PAR (r(181) = -0.30, p<0.001), with higher age indicating more anterior-dominant alpha. This relation remained significant after correcting for patients total CRS-R score. To account for the possible sex-based differences, we performed a second post-hoc analysis, investigating the sex-based differences in the diagnostic markers of all candidate features (see supplementary Figure S7). Most interestingly, the alpha PAR in non-anoxic patients showed a diverging effect between men and women. The alpha PAR showed a significant correlation with non-anoxic patients’ CRS-R score only in women, but not men. The analysis performed by Colombo et al. (2023) did not account for participants’ age and sex. Possible interaction effects should be further investigated to avoid age- or sex-based biases in the clinical assessment of consciousness.

### The EEG spectral exponent might have prognostic value for anoxic unresponsive patients

In addition to the diagnostic value for non-anoxic patients, this study suggests that the EEG spectral exponent has prognostic value for anoxic patients. Specifically, anoxic patients with a steeper spectral exponent had a higher probability for recovery of consciousness. A post-hoc analysis further confirmed a correlation between the spectral slope and anoxic patients’ quality of recovery (i.e., maximally achieved GOSE score within one year) (r(51)=-0.30, p<0.05), with a steeper spectral slope indicating a higher GOSE score and thus better recovery. Previous results provide evidence linking the flattening of the spectral exponent to higher levels of consciousness (Colombo et al. 2019; Lendner et al. 2020; Maschke, Duclos, Owen, et al. 2022). Our group previously demonstrated paradoxical effects on the spectral exponent and signal complexity following exposure to anesthesia (Maschke, Duclos, and Blain-Moraes 2022; Maschke, Duclos, Owen, et al. 2022), where overly-steep spectral exponents flattened in response to anesthesia. We therefore suggest that the EEG spectral exponent can be related to different underlying pathologies: 1) an over-steepening of the spectral exponent indicating altered excitation-inhibition balance; and 2) an over-flattening of the spectral exponent indicating global neuronal cell death and severity of injury in anoxic patients. A previous study investigating the prognostic value of the EEG spectral exponent in cardiac arrest patients did not find a significant difference between recovered and non-recovered patients (Alnes et al. 2021). However, outcome measures in this study might have been affected by the withdrawal of life sustaining treatment and the estimation of the spectral exponent while patients were sedated and in hypothermic treatment. More research needs to be done to further investigate the prognostic value of the spectral exponent for anoxic patients.

### Changes in signal complexity can be largely, but not exclusively, attributed to spectral properties

The analysis of spontaneous and evoked brain complexity has gained increasing attention as a marker of human consciousness. Reduced brain complexity and entropy of spontaneous EEG activity has been shown during anesthesia-induced unconsciousness (Bruhn et al. 2000; Zhang et al. 2001; Jordan et al. 2008; Schartner et al. 2015), sleep (Burioka et al. 2005; Mateos et al. 2018), epilepsy (Mateos et al. 2018) and in disorders of consciousness (Sarà and Pistoia 2010; Gosseries et al. 2011; Sitt et al. 2014; Stefan et al. 2018; Lei et al. 2022). In this study we demonstrated that the diagnostic value of LZC was largely attributable to changes in the spectral exponent. After correcting the signal complexity to a phase-shuffled signal with similar spectral properties (i.e., making LZC more independent from the spectral properties) diagnostic value of LZC weakened or vanished completely. However, disentangling signal complexity from the spectral exponent also revealed some diagnostic power of LZC for anoxic patients, which was not present in the classic LZC. We confirm our hypothesize that loss of signal complexity in lower levels of consciousness is epiphenomenal to changes in the spectral exponent and highlight the potential of metrics of signal complexity that rely less on the spectral exponent. LZC is only one measure from a multitude of available complexity and entropy measures (Lau et al. 2022); more research is needed to identify measures of signal complexity which are better suited for the assessment of unresponsive patients.

### None of the candidate EEG markers generalized to patients with other injuries

In this study, none of the candidate EEG markers showed diagnostic value for the group of patients who were unresponsive caused by other etiologies. One possible explication for this is the high heterogeneity of this group, which was composed of a variety etiologies and different mechanisms leading to alterations in consciousness. Our results indicate the importance of carefully using the term ‘markers of consciousness’ in the context of the evaluation of unresponsive patients. Previous research has pointed out that correlates of unresponsiveness are not solely markers of consciousness, but are also conflated with indexes of connectedness (Casey et al. 2024). In addition to level of consciousness and connectedness, EEG markers for the clinical assessment of consciousness are likewise influenced by type and severity of injury. Anoxic and non-anoxic brain injuries are highly heterogenous, the aim of identifying a ‘one size fits all’ marker of level of or capacity for consciousness is overly idealistic in a clinical context. Although it is meaningful to investigate markers of loss of consciousness in the healthy brain through different mechanisms that result in unconsciousness, those mechanisms do not necessarily translate to the injured brain. Instead, we want to point out the need for markers for consciousness and recovery which account for the type and severity of brain injury, and address interindividual and etiology-dependent differences. There is an unmet need for more clinically homogenous large datasets for the investigation of etiology-dependent clinical markers. As an alternative, the use of perturbation for patient-assessment overcomes the problem of individual differences by enabling within-subject analysis. More research is needed on personalized medical approaches for clinical assessment to overcome the limitation of interindividual heterogeneity yielding differences in spontaneous EEG markers.

### Limitations

The results of this study need to be interpreted in light of several limitations.

First, level of consciousness was assessed using the proxy of responsiveness (CRS-R score). However, the absence of responsiveness is not sufficient to claim the absence of consciousness (Sanders et al. 2012). To partially address this limitation, we conducted five variations of the diagnostic analysis: a basic diagnostic analysis (analysis 1-3); an analysis focused on patient responsiveness (analysis 4); and an analysis focused on the patient’s potential to recover (analysis 5). To maximally exclude confounding factors beyond the pure level and content of consciousness, we additionally analyzed the minimal contrast between UWS and MCS, MCS- and MCS+. Still, the diagnosis which relies on behavior alone is insufficient to correctly reflect the state of an unresponsive patient and should be ideally paired with evidence from functional brain imaging assessment (Naccache 2018).

Second, we combined traumatic brain injury and stroke into the group of non-anoxic patients. The same grouping has been performed in previous research (Colombo et al. 2023), in which they reported diverging effects between anoxic and non-anoxic patients but similar results within the group of non-anoxic patients (i.e., between stroke and TBI patients). In line with this, we performed analysis 1 (i.e., the correlation between candidate features and patients’ diagnosed level of consciousness) on TBI and stroke patients individually and provided the results in the supplementary material (see supplementary Figure S4). While similar trends were observed in the individual groups, the possibility to draw conclusions based on statistics is limited by the small sample size of 22 stroke patients. Despite similar cognitive recovery process (Castor and El Massioui 2018), TBI and stroke are still two distinct clinical conditions which should be treated and investigated independently. More research is needed to validate candidate EEG markers for their diagnostic and prognostic value for stroke and TBI patients independently.

Third, we only investigated space-averaged EEG features and did not account for the location and severity of injury. Investigating the diagnostic and prognostic value of specific brain regions’ EEG values would require accounting for individual characteristics of location and size of injury, which was not available in this dataset. Previous studies demonstrated a steepening of the spectral exponent in the stroke-affected hemisphere, compared to the healthy hemisphere (Lanzone et al. 2022). To partially address the spatial variability in EEG features, we performed a post-hoc analysis investigating the prognostic value of the standard deviation of the spectral exponent. More research is required to investigate spatial characteristics of EEG features for the prediction of outcome.

## 6 Conclusion

Identifying reliable markers of consciousness is fundamental for the assessment unresponsive patients. Alpha power has long been considered a reliable and generalizable clinical index of consciousness; our results support the claim that alpha power is an etiology-specific marker, which has higher diagnostic value for anoxic patients. In addition, we highlight the diagnostic value of the spectral exponent for the assessment of non-anoxic patients and point out the strong dependence between complexity and the spectral exponent. Our results demonstrate the importance of considering etiologies independently when investigating markers for capacity for consciousness. We demonstrated clinical limitations of current EEG markers of consciousness which are not generalizable across different brain pathologies. Moving forward, we encourage the development of within-subject approaches to address the limitation of interindividual heterogeneity.

## Data Availability

The participants of this study did not give consent for their data to be shared publicly. Due to the sensitive nature of the research EEG data is not publicly available. Derived data supporting the findings of this study are available upon reasonable request to the authors.

## 7. Acknowledgments

SBM is supported by the Canada Research Chairs Program (Tier II). CM is funded through Fonds de recherche du Québec – Santé (FRQS). This research is supported in part by the FRQNT Strategic Clusters Program (2020-RS4-265502 - Centre UNIQUE - Union Neurosciences & Artificial Intelligence – Quebec). This study was funded through an NSERC Discovery Grant (RGPIN-2023-03619) and a France Canada Research Fund New Research Collaborations Program. This research was undertaken thanks in part to funding from the Canada First Research Excellence Fund and Fonds de recherche du Québec, awarded to the Healthy Brains, Healthy Lives initiative at McGill University. This study was partly funded by the European Union (ERA PerMed JTC2019 “PerBrain”, grant to JDS) and FLAG-ERA project ModelDXConsciousness. DM received individual funding from Ecole Doctorale Frontières de l’Innovation en Recherche et Education–Programme Bettencourt.

