## supplementary for "The role of etiology in the identification of clinical markers of consciousness: comparing EEG alpha power, complexity, and spectral exponent"

Content:

- Supplementary Figure S1- S7
- Supplementary Table S1- S5

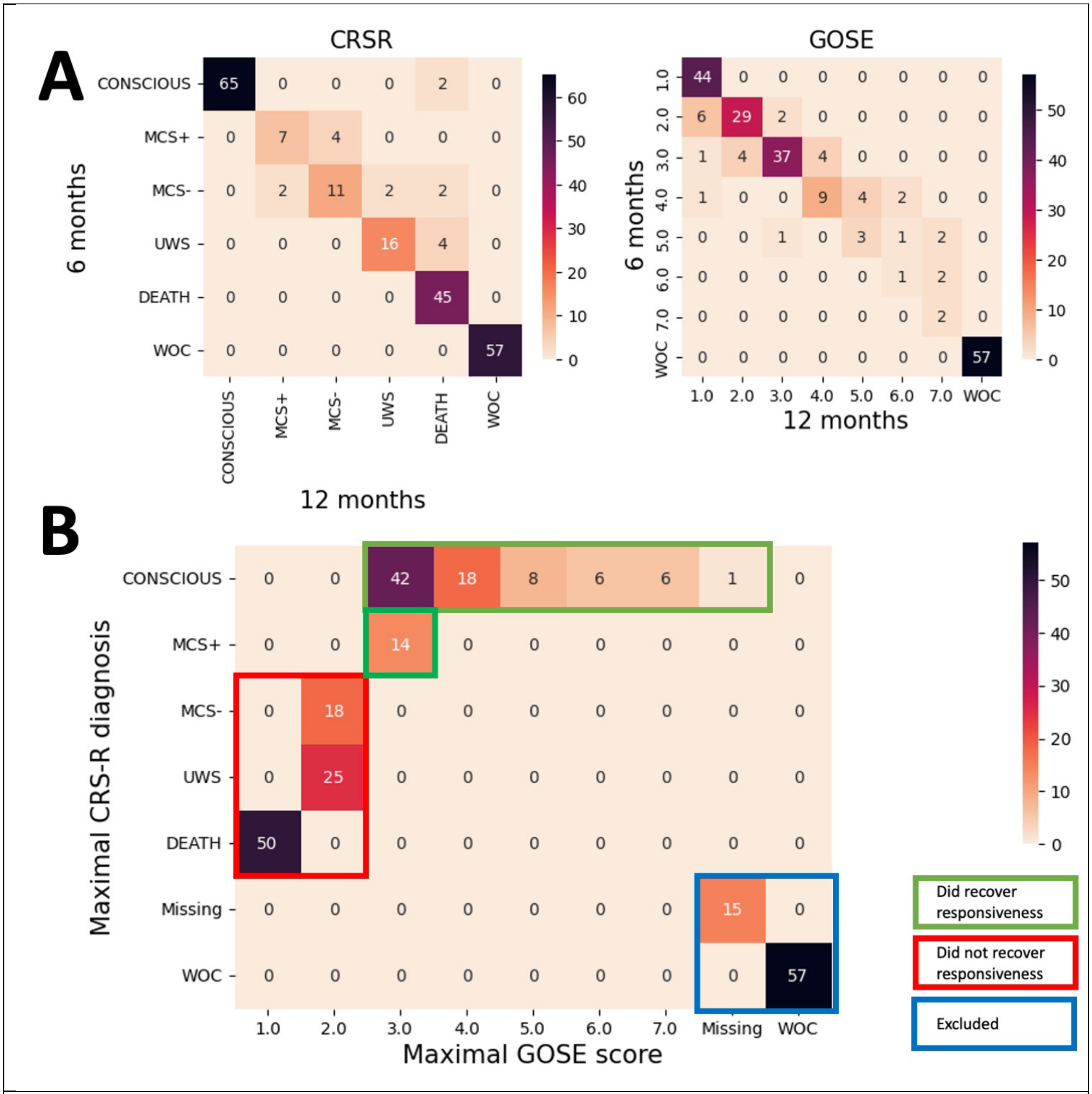


**Figure S1.** Distribution of outcome values assessed by Coma Recovery Scale Revised (CRS-R) and Glasgow Outcome Scale Extended (GOSE). **A** Outcome based on CRS-R score (left) and GOSE (right) at 6 and 12 months. **B** Grouping of patients who did (green) or did not (red) recover responsiveness within 12 months post injury. Patients whose reason of death was withdrawal of care (WOC) were excluded from the prognostic analysis.
CRSR: Coma Recovery Scale Revised, GOSE: Glasgow outcome scale extended, UWS: Unresponsive Wakefulness Syndrome, MCS: Minimally Conscious State

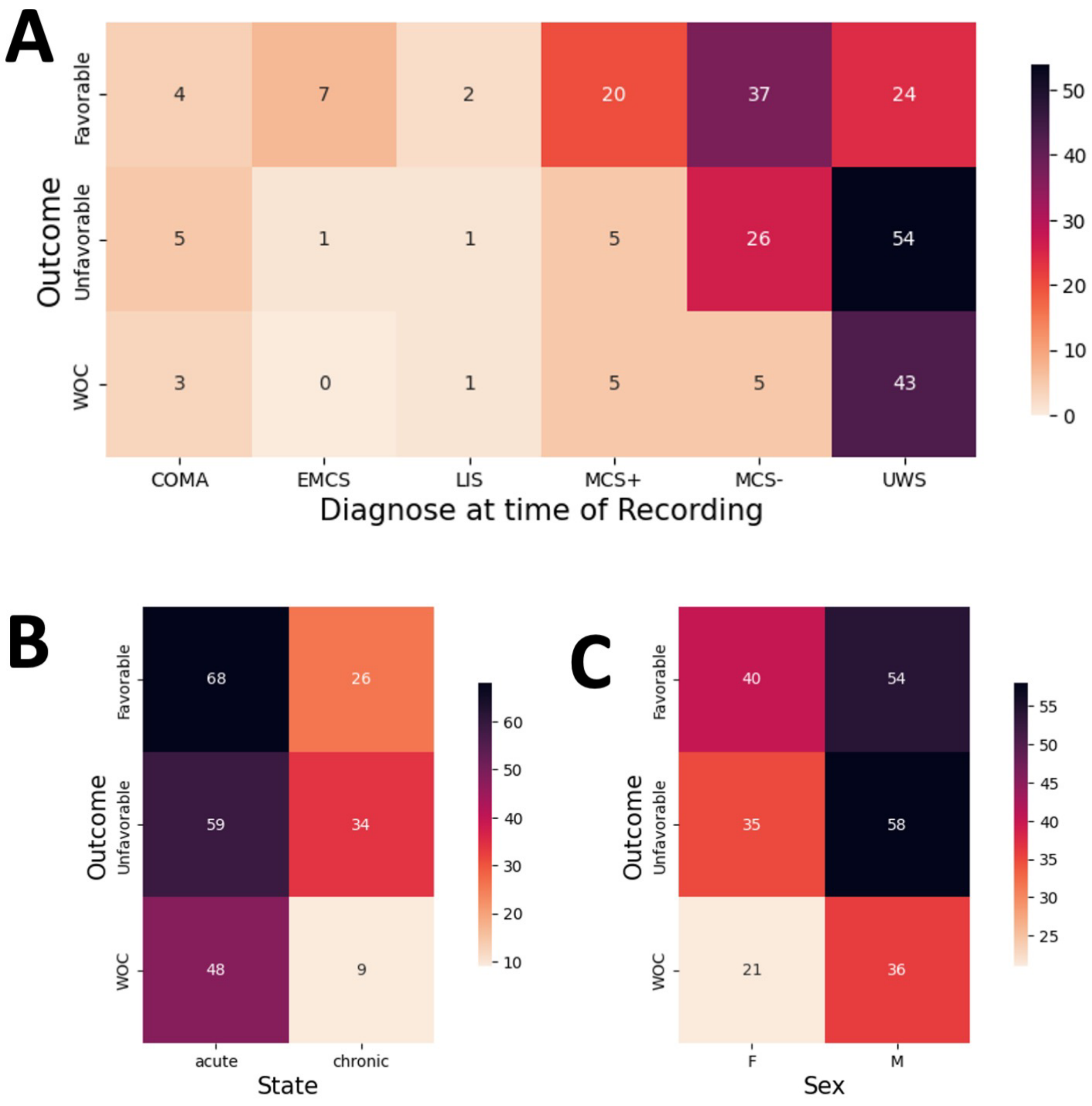


**Figure S2.** Outcome demographics. **A:** Distribution of outcome values split by patient diagnose at time of recording. **B:** Distribution of outcome values split by medical state of the patient. **C:** Distribution of outcome values split by patients' sex. Patients whose reason of death was withdrawal of care (WOC) were excluded from the prognostic analysis.
UWS: Unresponsive Wakefulness Syndrome, MCS: Minimally Conscious State, EMCS: Emergence, LIS: Locked in Syndrome

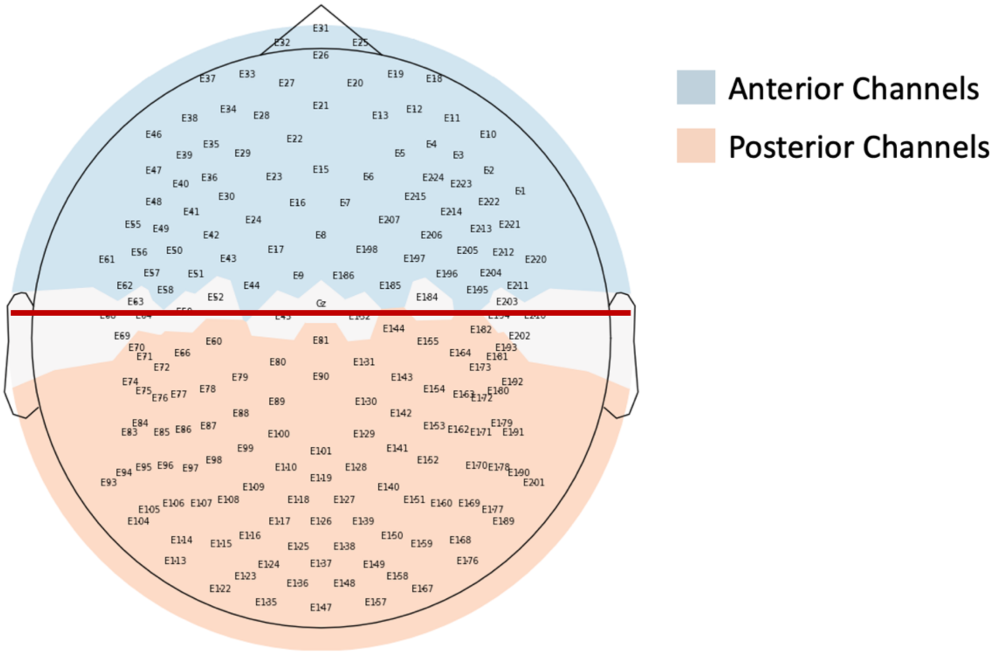


**Figure S3.** Topographic distribution of anterior and posterior channels.

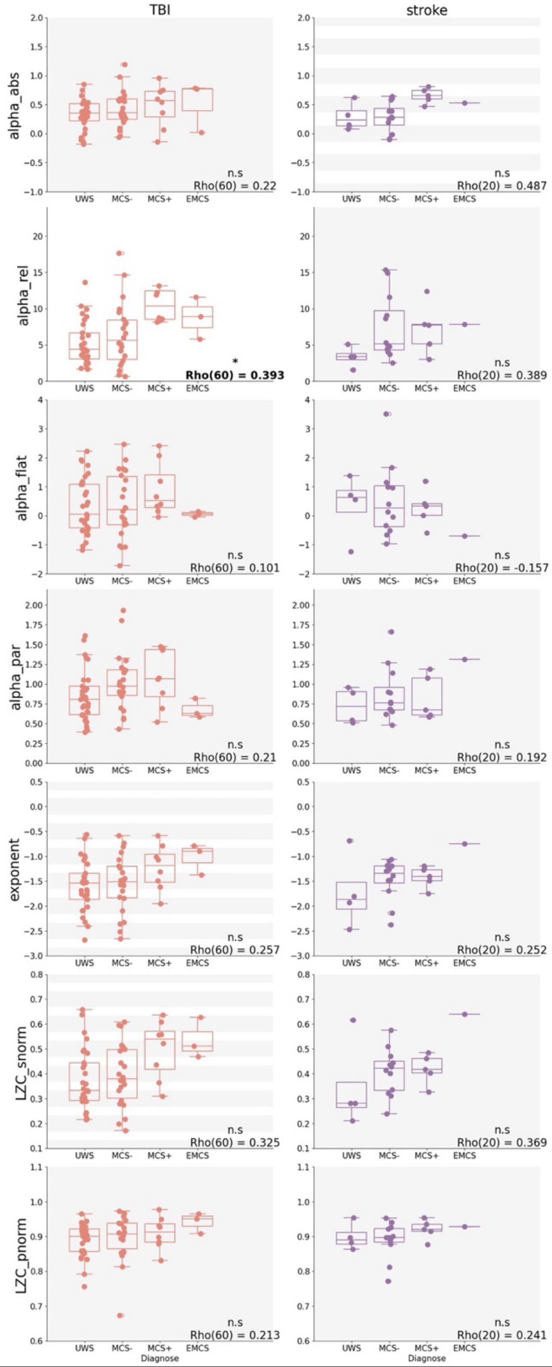


**Figure S4. Correlation between candidate features and non-anoxic patients' diagnosed level of consciousness, split by etiologies into traumatic brain injury and stroke.** Spearman rank correlation value and degrees of freedom are indicated for each correlation. Significant correlations (p<0.05 after correction) are written in bold with white background. A grey background indicates non-significant correlation. A background with grey and white lines indicates loss of significance after p-value correction.

UWS: Unresponsive Wakefulness Syndrome, MCS: Minimally Conscious State, EMCS: Emergence, TBI: Traumatic brain injury

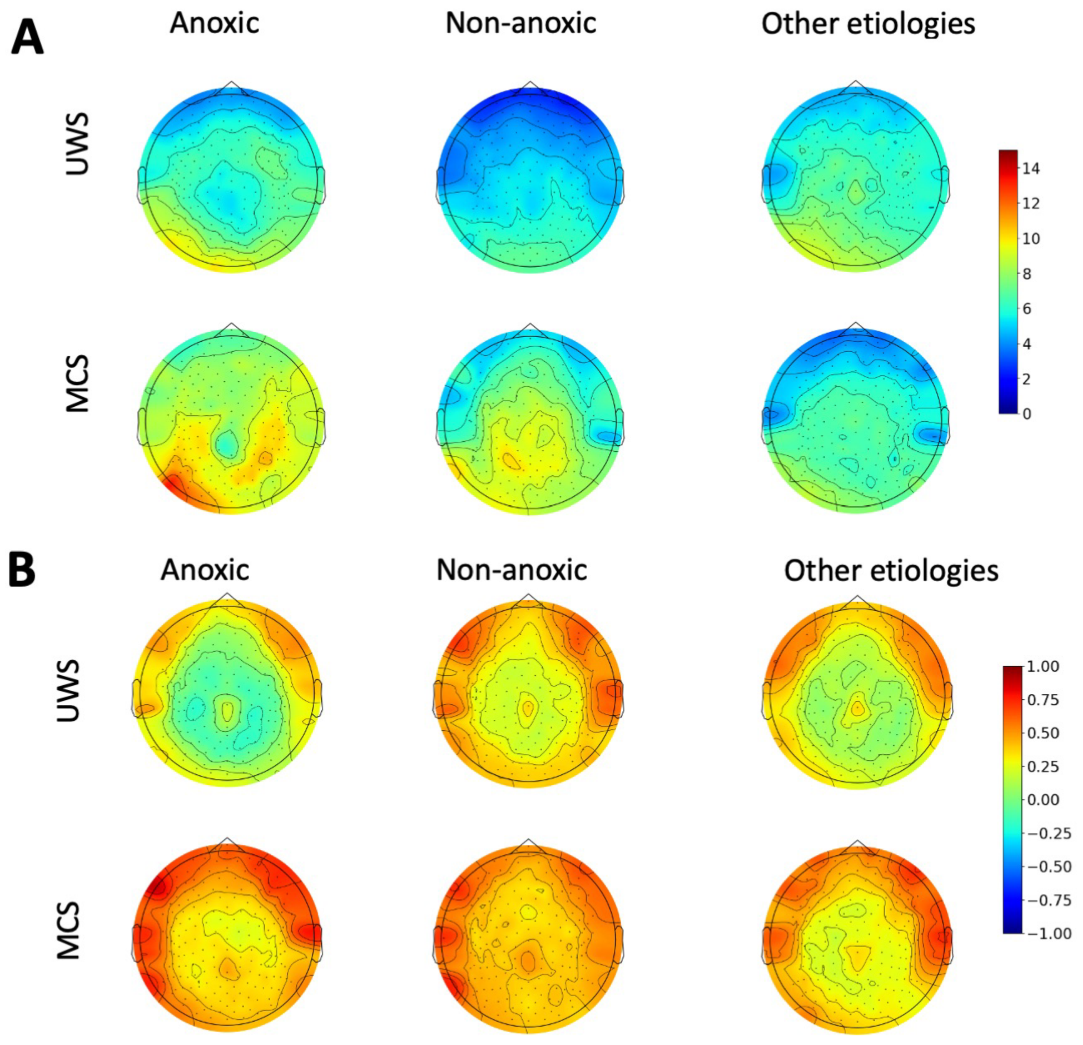


**Figure S5: Relative and absolute alpha power in different etiologies and levels of consciousness.**

Topographic visualization of (A) relative alpha power and (B) absolute alpha power between patients in a UWS and MCS, split by etiology. Topographic maps represent group-averaged power. UWS: Unresponsive Wakefulness Syndrome, MCS: Minimally Conscious State

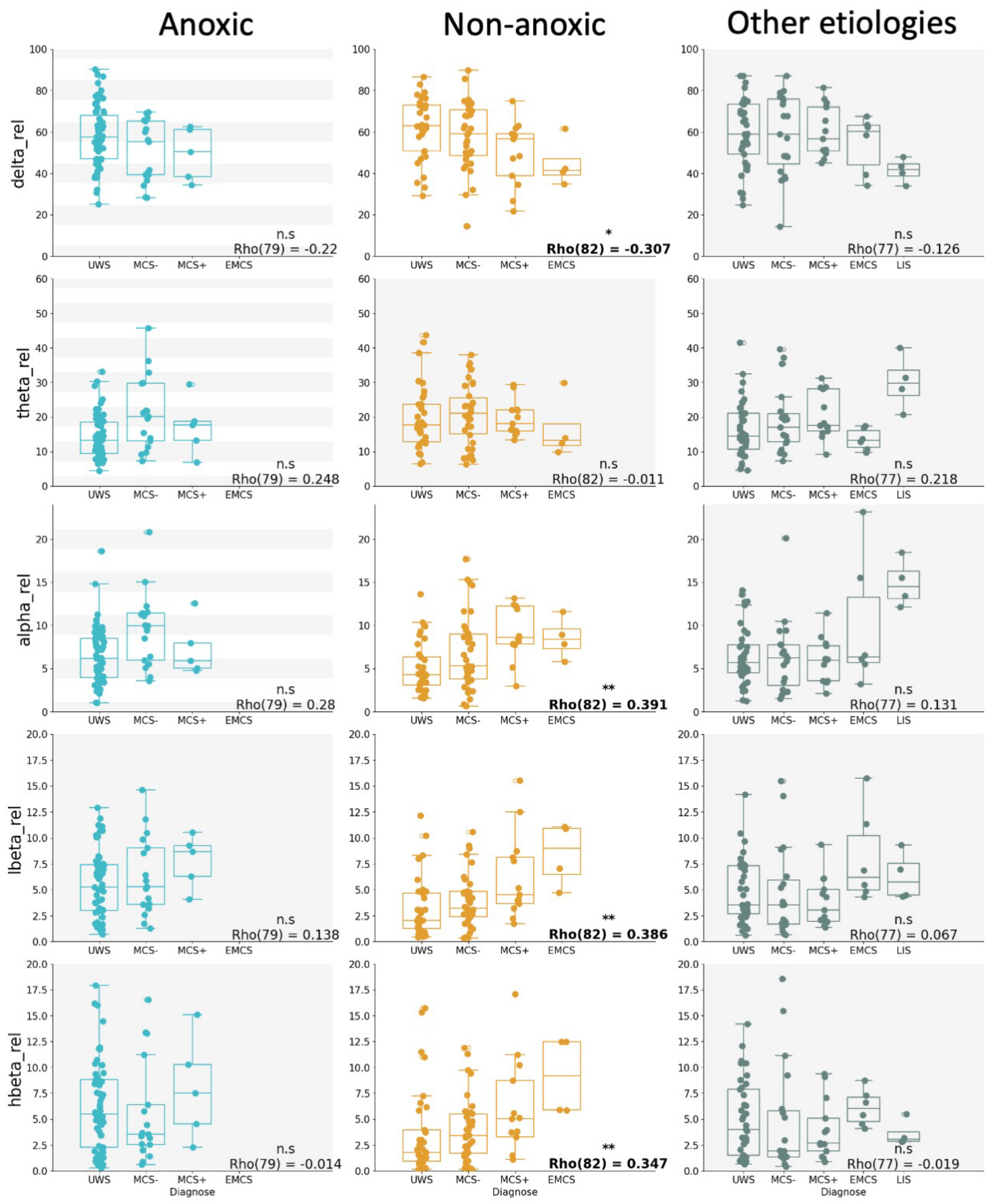


**Figure S6: Correlation between levels of consciousness and relative power in different etiologies and frequency bands.**

Spearman rank correlation value and degrees of freedom are indicated for each correlation. Significant correlations (p<0.05 after correction) are written in bold with white background. Corrected significance values are indicated above, with * indicating p< 0.05, ** being p < 0.01, *** being p< 0.001 and n.s. indicating no significance. A grey background indicates non-significant correlation. A background with grey and white lines indicates loss of significance after p-value correction. Each dot corresponds to an individual patient.

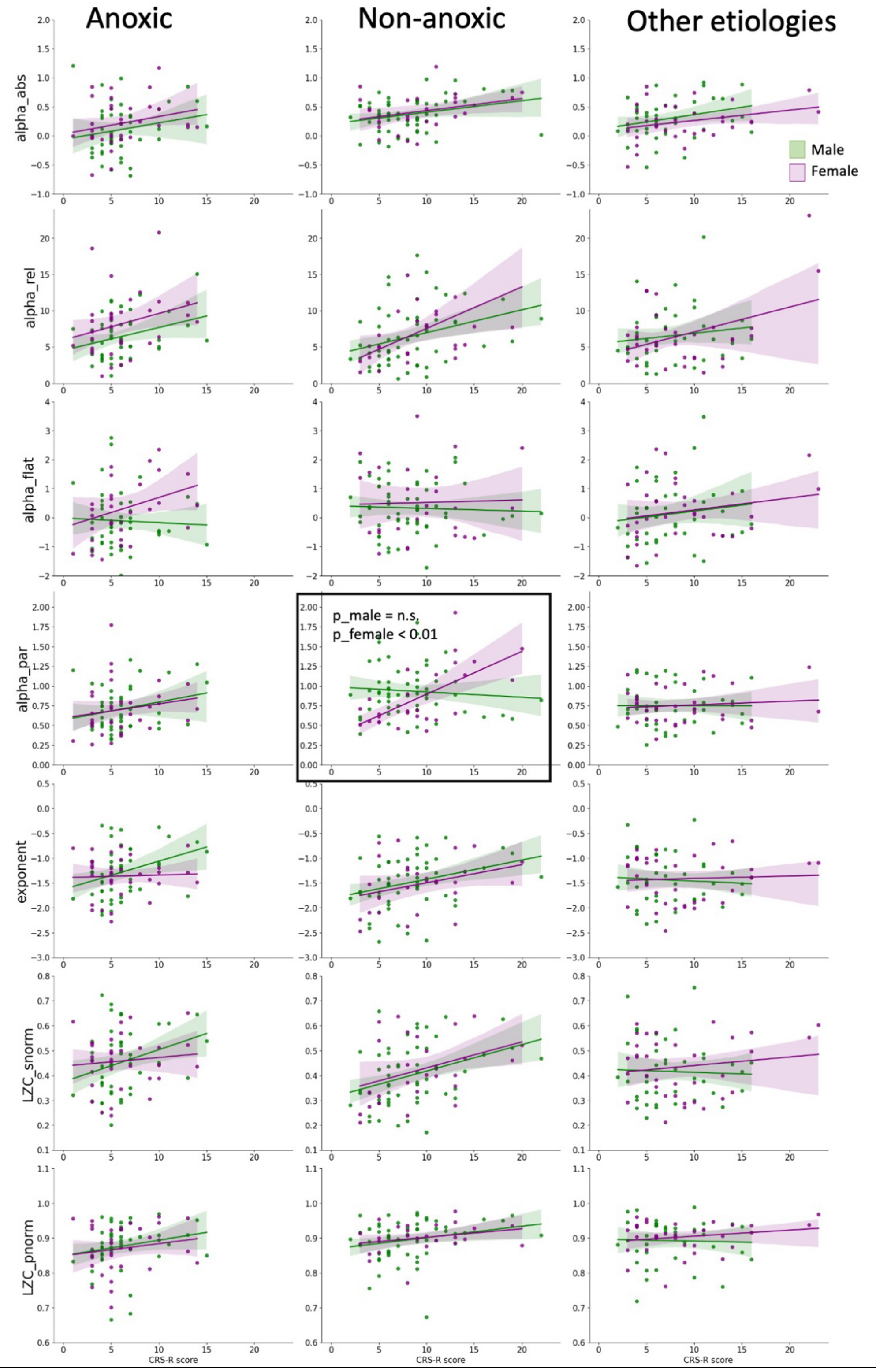


**Figure S7: Correlation between patients CRSR score and candidate features, split by etiology and patient sex.**

Green dots represent male participants, purple dots female participants.

CRSR: Coma Recovery Scale Revised

**Table S1. Statistical values for analysis 1:** Correlation between candidate measures and patient's diagnosed level of consciousness. dof: degrees of freedom; PAR: Posterior-Anterior ratio; LZC: Lempel-Ziv Complexity; rho: spearman rho; snorm: shuffle-normalized; pnorm: phase-normalized.

| **All participants** | rho | dof | p raw | p corrected |
|---|---|---|---|---|
| Alpha absolute | 0.305 | 238 | **< 0.001** | **< 0.001** |
| Alpha relative | 0.188 | 238 | **0.0035** | **0.0245** |
| Alpha flat | 0.161 | 238 | **0.0127** | 0.0889 |
| Alpha PAR | 0.19 | 238 | 0.0032 | **0.0224** |
| Exponent | 0.052 | 238 | 0.4267 | 1 |
| LZC snorm | 0.087 | 238 | 0.1798 | 1 |
| LZC pnorm | 0.186 | 238 | **0.004** | **0.028** |

| **Anoxic** | rho | dof | p raw | p corrected |
|---|---|---|---|---|
| Alpha absolute | 0.379 | 79 | **< 0.001** | **0.0035** |
| Alpha relative | 0.28 | 79 | **0.012** | 0.084 |
| Alpha flat | 0.312 | 79 | **0.0048** | **0.0336** |
| Alpha PAR | 0.161 | 79 | 0.1537 | 1 |
| Exponent | 0.016 | 79 | 0.8876 | 1 |
| LZC snorm | 0.035 | 79 | 0.7612 | 1 |
| LZC pnorm | 0.215 | 79 | 0.0555 | 0.3885 |

| **Non-anoxic** | rho | dof | p_raw | p_corr |
|---|---|---|---|---|
| Alpha absolute | 0.262 | 82 | **0.0167** | 0.1169 |
| Alpha relative | 0.391 | 82 | **< 0.001** | **0.0021** |
| Alpha flat | 0.022 | 82 | 0.8464 | 1 |
| Alpha PAR | 0.197 | 82 | 0.0737 | 0.5159 |
| Exponent | 0.281 | 82 | **0.01** | 0.07 |
| LZC snorm | 0.341 | 82 | **0.0016** | **0.0112** |
| LZC pnorm | 0.22 | 82 | **0.046** | 0.322 |

| **Other etiologies** | rho | dof | p_raw | p_corr |
|---|---|---|---|---|
| Alpha absolute | 0.209 | 77 | 0.0677 | 0.4739 |
| Alpha relative | 0.131 | 77 | 0.2557 | 1 |
| Alpha flat | 0.172 | 77 | 0.1337 | 0.9359 |
| Alpha PAR | 0.13 | 77 | 0.2613 | 1 |
| Exponent | -0.052 | 77 | 0.6553 | 1 |
| LZC snorm | 0.051 | 77 | 0.6589 | 1 |
| LZC pnorm | 0.126 | 77 | 0.274 | 1 |

**Table S2. Statistical values for analysis 2:** Correlation between candidate measures and patient's CRSR score. dof: degrees of freedom; PAR: Posterior-Anterior ratio; LZC: Lempel-Ziv Complexity; rho: spearman rho; snorm: shuffle-normalized; pnorm: phase-normalized.

| **All participants** | rho | dof | p_raw | p_corr |
|---|---|---|---|---|
| Alpha absolute | 0.276 | 238 | **< 0.001** | **< 0.001** |
| Alpha relative | 0.245 | 238 | **< 0.001** | **< 0.001** |
| Alpha flat | 0.129 | 238 | **0.046** | 0.322 |
| Alpha PAR | 0.192 | 238 | **0.0029** | **0.0203** |
| Exponent | 0.113 | 238 | 0.0819 | 0.5733 |
| LZC snorm | 0.147 | 238 | **0.023** | 0.161 |
| LZC pnorm | 0.211 | 238 | **0.001** | **0.007** |
| **Anoxic** | rho | dof | p_raw | p_corr |
| Alpha absolute | 0.192 | 79 | 0.0873 | 0.6111 |
| Alpha relative | 0.267 | 79 | **0.0167** | 0.1169 |
| Alpha flat | 0.123 | 79 | 0.2767 | 1 |
| Alpha PAR | 0.199 | 79 | 0.0764 | 0.5348 |
| Exponent | 0.209 | 79 | 0.0631 | 0.4417 |
| LZC snorm | 0.209 | 79 | 0.0627 | 0.4389 |
| LZC pnorm | 0.234 | 79 | **0.0369** | 0.2583 |
| **Non-anoxic** | rho | dof | p_raw | p_corr |
| Alpha absolute | 0.306 | 82 | **0.005** | **0.035** |
| Alpha relative | 0.434 | 82 | **< 0.001** | **< 0.001** |
| Alpha flat | -0.021 | 82 | 0.8476 | 1 |
| Alpha PAR | 0.213 | 82 | 0.0531 | 0.3717 |
| Exponent | 0.326 | 82 | **0.0027** | **0.0189** |
| LZC snorm | 0.391 | 82 | **< 0.001** | **0.0021** |
| LZC pnorm | 0.278 | 82 | **0.0111** | 0.0777 |
| **Other etiologies** | rho | dof | p_raw | p_corr |
| Alpha absolute | 0.222 | 77 | 0.0569 | 0.3983 |
| Alpha relative | 0.115 | 77 | 0.3286 | 1 |
| Alpha flat | 0.185 | 77 | 0.1154 | 0.8078 |
| Alpha PAR | -0.041 | 77 | 0.7259 | 1 |
| Exponent | -0.09 | 77 | 0.4476 | 1 |
| LZC snorm | -0.025 | 77 | 0.8294 | 1 |
| LZC pnorm | 0.012 | 77 | 0.9219 | 1 |

**Table S3. Statistical values for analysis 3:** Diagnostic value of candidate measures to distinguish patients in an Unresponsive Wakefulness Syndrome from patients with higher levels of consciousness (i.e. Minimally conscious state and emergence). PAR: Posterior-Anterior ratio; LZC: Lempel-Ziv Complexity; snorm: shuffle-normalized; pnorm: phase-normalized.

| **All participants** | U | p_raw | p_corr |
|---|---|---|---|
| Alpha absolute | 4667 | **< 0.001** | **< 0.001** |
| Alpha relative | 5743 | **0.0083** | 0.0581 |
| Alpha flat | 5597 | **0.0036** | **0.0252** |
| Alpha PAR | 5405 | **0.0011** | **0.0077** |
| Exponent | 7108 | 0.9242 | 1 |
| LZC snorm | 6869 | 0.5887 | 1 |
| LZC pnorm | 5871 | **0.0163** | 0.1141 |

| **Anoxic** | U | p_raw | p_corr |
|---|---|---|---|
| Alpha absolute | 321 | **< 0.001** | **0.0035** |
| Alpha relative | 396 | **0.0073** | 0.0511 |
| Alpha flat | 363 | **0.0024** | **0.0168** |
| Alpha PAR | 497 | 0.1077 | 0.7539 |
| Exponent | 647 | 0.9873 | 1 |
| LZC snorm | 630 | 0.8443 | 1 |
| LZC pnorm | 474 | 0.0639 | 0.4473 |

| **Non-anoxic** | U | p_raw | p_corr |
|---|---|---|---|
| Alpha absolute | 655 | 0.0885 | 0.6195 |
| Alpha relative | 522 | **0.0035** | **0.0245** |
| Alpha flat | 805 | 0.7416 | 1 |
| Alpha PAR | 655 | 0.0885 | 0.6195 |
| Exponent | 639 | 0.0643 | 0.4501 |
| LZC snorm | 597 | **0.0254** | 0.1778 |
| LZC pnorm | 683 | 0.1479 | 1 |

| **Other etiologies** | U | p_raw | p_corr |
|---|---|---|---|
| Alpha absolute | 557 | 0.1231 | 0.8617 |
| Alpha relative | 700 | 0.9789 | 1 |
| Alpha flat | 620 | 0.382 | 1 |
| Alpha PAR | 640 | 0.5078 | 1 |
| Exponent | 779 | 0.4237 | 1 |
| LZC snorm | 742 | 0.6833 | 1 |
| LZC pnorm | 710 | 0.9451 | 1 |

**Table S4. Statistical values for analysis 4:** Diagnostic value of candidate measures to distinguish unresponsive patients (i.e. unresponsive wakefulness syndrome and minimally conscious state -) from patients with some level of responsiveness (i.e. Minimally conscious state+ and emergence). PAR: Posterior-Anterior ratio; LZC: Lempel-Ziv Complexity; snorm: shuffle-normalized; pnorm: phase-normalized.

| **All participants** | U | p_raw | p_corr |
|---|---|---|---|
| Alpha absolute | 2883 | **0.0031** | **0.0217** |
| Alpha relative | 3153 | **0.0222** | 0.1554 |
| Alpha flat | 3839 | 0.5533 | 1 |
| Alpha PAR | 3673 | 0.3159 | 1 |
| Exponent | 3328 | 0.0636 | 0.4452 |
| LZC snorm | 3096 | **0.0152** | 0.1064 |
| LZC pnorm | 3042 | **0.0104** | 0.0728 |

| **Anoxic** | U | p_raw | p_corr |
|---|---|---|---|
| Alpha absolute | 138 | 0.3224 | 1 |
| Alpha relative | 185 | 0.9318 | 1 |
| Alpha flat | 198 | 0.8866 | 1 |
| Alpha PAR | 208 | 0.739 | 1 |
| Exponent | 136 | 0.3036 | 1 |
| LZC snorm | 139 | 0.3321 | 1 |
| LZC pnorm | 117 | 0.1594 | 1 |

| **Non-anoxic** | U | p_raw | p_corr |
|---|---|---|---|
| Alpha absolute | 320 | **0.0056** | **0.0392** |
| Alpha relative | 269 | **0.0008** | **0.0056** |
| Alpha flat | 518 | 0.5702 | 1 |
| Alpha PAR | 525 | 0.6242 | 1 |
| Exponent | 344 | **0.0122** | 0.0854 |
| LZC snorm | 296 | **0.0024** | **0.0168** |
| LZC pnorm | 390 | **0.0463** | 0.3241 |

| **Other etiologies** | U | p_raw | p_corr |
|---|---|---|---|
| Alpha absolute | 451 | 0.3268 | 1 |
| Alpha relative | 509 | 0.784 | 1 |
| Alpha flat | 532 | 1 | 1 |
| Alpha PAR | 492 | 0.6304 | 1 |
| Exponent | 501 | 0.7102 | 1 |
| LZC snorm | 461 | 0.3904 | 1 |
| LZC pnorm | 498 | 0.6832 | 1 |

**Table S5. Statistical values for analysis 5:** Prognostic value of candidate measures to distinguish patients who recovered responsiveness from patients with an unfavorable outcome. PAR: Posterior-Anterior ratio; LZC: Lempel-Ziv Complexity; snorm: shuffle-normalized; pnorm: phase-normalized.

| **All participants** | U | p_raw | p_corr |
|---|---|---|---|
| Alpha absolute | 4538 | 0.3969 | 1 |
| Alpha relative | 3847 | 0.2877 | 1 |
| Alpha flat | 4611 | 0.2941 | 1 |
| Alpha PAR | 4208 | 0.9492 | 1 |
| Exponent | 3215 | **0.0049** | **0.0343** |
| LZC snorm | 3434 | **0.0274** | 0.1918 |
| LZC pnorm | 3870 | 0.3176 | 1 |

| **Anoxic** | U | p_raw | p_corr |
|---|---|---|---|
| Alpha absolute | 378 | 0.2405 | 1 |
| Alpha relative | 315 | 1 | 1 |
| Alpha flat | 414 | 0.0643 | 0.4501 |
| Alpha PAR | 340 | 0.6454 | 1 |
| Exponent | 175 | **0.0088** | 0.0616 |
| LZC snorm | 190 | **0.0194** | 0.1358 |
| LZC pnorm | 306 | 0.8732 | 1 |

| **Non-anoxic** | U | p_raw | p_corr |
|---|---|---|---|
| Alpha absolute | 700 | 0.5883 | 1 |
| Alpha relative | 601 | 0.5807 | 1 |
| Alpha flat | 727 | 0.3995 | 1 |
| Alpha PAR | 659 | 0.9333 | 1 |
| Exponent | 541 | 0.2217 | 1 |
| LZC snorm | 545 | 0.239 | 1 |
| LZC pnorm | 504 | 0.1021 | 0.7147 |

| **Other etiologies** | U | p_raw | p_corr |
|---|---|---|---|
| Alpha absolute | 357 | 0.3417 | 1 |
| Alpha relative | 377 | 0.5227 | 1 |
| Alpha flat | 362 | 0.3827 | 1 |
| Alpha PAR | 298 | 0.0614 | 0.4298 |
| Exponent | 383 | 0.5853 | 1 |
| LZC snorm | 420 | 0.9876 | 1 |
| LZC pnorm | 370 | 0.4543 | 1 |
